# Where does healthcare worker time go? Evidence from a time-and-motion study in Malawi

**DOI:** 10.64898/2026.05.04.26352396

**Authors:** Bingling She, Precious Chitsulo, Joseph H Collins, Watipaso Mulwafu, Emmanuel Mnjowe, Sangeeta Bhatia, Tara D Mangal, Sebastian Mboma, Sakshi Mohan, Margherita Molaro, Pemphero Norah Mphamba, Rachel E. Murray-Waston, Andrew Phillips, Paul Revill, Mariana Suarez, Victor Mwapasa, Dominic Nkhoma, Joseph Mfutso-Bengo, Timothy B Hallett, Wiktoria Tafesse, Tim Colbourn

**Author notes:** Corresponding author. School of Public Health, Imperial College London, London W12 0BZ, UK. Joint-last authors (Wiktoria Tafesse, Tim Colbourn).

## Abstract

Low- and middle-income countries face critical shortages of healthcare workers (HCWs) and funding for human resources for health (HRH), while patients often receive less care time than expected. Understanding how the existing workforce capacity is used is therefore essential for improving health system performance in resource-constrained settings. We examined HCW time-use patterns in Malawi using data from a time-and-motion study conducted between January and May 2024, which recorded activities across multiple cadres, days, and representative health facilities in the healthcare system. Across cadres, median daily working time, including breaks, was 7.35 hours (IQR 4.40-8.35), approximately 1.65 hours below the typical contracted schedule. HCWs spent most time on direct patient care: 2.82 hours per day (IQR 1.89-3.97), accounting for 48% of total working time (IQR 30%-67%). Administrative tasks accounted for 0.30 hours (IQR 0.00-1.23; 5.21%, IQR 0%-18%) and break time remained consistent with the contracted expectations at 1.25 hours (IQR 0.00-2.12; 18%, IQR 0%-28%). Unallocated time, defined as time neither work-related nor recorded as breaks, was 0.72 hours (IQR 0.02-1.92; 12%, IQR 0%-29%), mainly attributed to the absence of patients based on available information. Median patient load was 21 per staff member per day in outpatient care (IQR 12-35), 12 in inpatient care (IQR 7-18), and 14 in emergency care (IQR 10-23), with median time per patient of 3 (IQR 1.0-6.5), 6 (IQR 2.5-14), and 10 (IQR 5-20) minutes, respectively. These measures, particularly time per patient, vary by cadre, facility type, facility ownership, region, and service area. The findings present a first system-wide picture of HCW time use in a low-income setting and can inform health systems planning. The gap between contracted and actual working time and unallocated time suggests scope to improve workforce utilisation, while high patient loads highlight the need for sustained HRH investment and workforce expansion.

**Key Messages:**

- In low- and middle-income countries with persistent health workforce and human resources for health (HRH) funding constraints, it is essential to understand how healthcare worker (HCW) time is utilised in practice to identify opportunities to improve service delivery and overall health system performance.
- Based on a time-and-motion study in Malawi health system, we observed that HCWs worked a median of 7.35 hours per day (including breaks), below the typical contracted schedule. Although most working time was devoted to direct patient care, the patient-facing time was limited relative to high patient loads, with short service time per patient, particularly in outpatient settings. The time-use patterns also varied across HCW cadres, facility types, regions, facility ownership, and service areas.
- Workforce planning should address both utilisation and capacity: reducing avoidable unallocated time may improve efficiency, but high patient loads and short service time per patient indicate that sustained HRH investment and workforce expansion remain essential.

## Introduction

In low- and middle-income countries (LMICs), health care demand is rising due to demographic and epidemiological transitions, particularly population growth and an increasing burden of non-communicable diseases, alongside broader socioeconomic changes such as ongoing urbanisation contributing to shifts in service utilisation patterns through improved access and health awareness (Kawale, Pagliari and Grant, 2019; Molaro *et al*., 2025; Choi *et al*., 2016; Gagnon-Dufresne *et al*., 2023). However, health service provision is limited by a shortage of health care workers (HCWs) due to lack of funding (Muula, 2023), among other health system constraints. This situation may be worsened in the near future because of recent donor funding cuts (Molaro *et al*., 2025). In such highly resource-constrained settings, it is crucial to understand current HCW time use patterns, identify possible inefficiencies, and provide empirical evidence to guide better use of available HCWs time that benefits health care provision and population health outcomes (Muho, Peshkatari and Wyss, 2022; Muho *et al*., 2025).

Observation-based time-and-motion studies (TMS) offer a valuable approach to understanding how HCWs allocate their time across different tasks and services (Muho *et al*., 2025; Kalne and Mehendale, 2022; Lopetegui *et al*., 2014). However, most previous studies, including those conducted in Malawi, have focused on single cadres such as community health workers, doctors, or nurses, in a limited number of facilities, or on specific diseases or health programs at only community or primary care level (Muho *et al*., 2025; Aron *et al*., 2023; Chinkhumba *et al*., 2022; Muho, Peshkatari and Wyss, 2022; Brar *et al*., 2021; Chebolu-Subramanian *et al*., 2019; Singh *et al*., 2018; Bonfim *et al*., 2016; Tilahun *et al*., 2017; Gardner *et al*., 2010; Gottschalk and Flocke, 2005; Pizziferri *et al*., 2005; Sessions *et al*., 2019; Jafry *et al*., 2016). In addition, there is limited evidence on the extent of unallocated hours - periods when staff are on site but not engaged in direct patient care, administration, or other work-related tasks, nor having permitted breaks - despite their implications for workforce efficiency (Muho, Peshkatari and Wyss, 2022; Bryant and ESSOMBA, 1995). Other studies use HCW self-reported information to assess time use, which however share the narrow focus described above and suffer from recall and social desirability biases (Garg *et al*., 2022; Jain *et al*., 2020; Wani *et al*., 2021; Anskär *et al*., 2018; Castellani *et al*., 2016; Burke *et al*., 2000). These indicate an evidence gap in a comprehensive understanding of time use across the broader health workforce and routine service delivery, i.e., multiple HCW cadres, different levels of care, and various service areas that can better represent health systems.

To address these gaps, we conducted a TMS of HCWs across multiple cadres, facility types at all levels of care, facility ownerships, regions, rural/urban areas, and various service areas in Malawi health care system from January to May 2024. Using the data collected, this study describes the HCW daily working time, time allocation across direct patient care, administration, and other tasks, the patient load (i.e., the number of patients seen), and time per patient, by these characteristics. By providing an empirical and holistic picture of HCW time use, we aim to inform evidence-based workforce and health systems planning, and improve the alignment of health worker time use, service delivery, facility management and health system resilience in Malawi and similar settings.

## Methods

### Malawi health care system

Health services in Malawi are delivered through a four-level system: the community level, consisting mainly of Health Posts and Outreach Clinics; the primary level, including Health Centres and Community/Rural Hospitals; the secondary level, comprising District Hospitals and faith-based hospitals with similar capacity; and the tertiary (regional) level, represented by central hospitals (She *et al*., 2024; Government of Malawi, 2018). As of 2024, the system comprised 1601 health facilities, based on Master Health Facility Register of Malawi, and 34,486 HCWs, according to Ministry of Health (MoH) records. The majority of health services are provided by public facilities managed by MoH, and non-profit non-governmental faith-based facilities managed by Christian Health Association of Malawi (CHAM), which are publicly supported and integrated into the public health system; less than 3% of services are provided by other non-governmental organisations including private for profit facilities (She *et al*., 2024; Government of Malawi, 2018; Government of Malawi, 2017; Varela *et al*., 2019). The official contracted working time for HCWs in the public sector is up to 48 hours per week according to Malawi Government Employment Act document, typically around 9 hours per day (07:30–16:30, including a 1-hour break), although this varies in practice by cadre, shift type (day, night, on-call, 24-hour), and facility-level management.

### Time-and-Motion study

Facilities in the 2023 Master Health Facility Register were first stratified by ownership (MoH and CHAM, then by rural/urban location, and subsequently by catchment population size (above or below the national average). Within each stratum, facilities were randomly selected via a random number generator in Microsoft Excel. Using this approach, 24 primary- and secondary-level facilities including Health Centres, Community/Rural Hospitals, and District Hospitals were sampled. All 5 tertiary hospitals (4 Central Hospitals and Zomba Mental Hospital) were additionally included to capture insights across levels of care. See Table A.1 in the Appendix for the characteristics of all selected facilities.

In every selected health facility, two trained data enumerators with clinical backgrounds randomly selected at least 6 HCWs (except Health Centres due to fewer HCWs) from the facility duty roster via a random number generator in Microsoft Excel, making sure a representative sample of mixed cadres including doctors, nurses, laboratory technicians, pharmacists, and others. There was also oversampling to include at least one doctor in smaller facilities. Without prior announcement of the start of HCW observation, each data enumerator followed at least 3 HCWs, one at a time, for 3 consecutive day shifts or for one night shift. Using the data-collection tool KoboToolbox (Kobo team), they recorded information of all activities performed by the HCWs including the activity labels, the start and end times (captured to the nearest minute), the service areas (i.e., clinics/wards/departments in the facility), and the health condition of any patients attended to. Each new patient seen by a HCW per shift was recorded and given a patient number. The activity labels, service areas, and patient health conditions were selected from standardised lists (with in total 127 activity labels, 21 health conditions, and 44 service areas) that were developed in collaboration with Malawian clinical experts (Nkhoma *et al*., 2025a; Nkhoma *et al*., 2025b), adapted to the existing literature (Aron *et al*., 2023).

The whole observation period at Health Centres lasted 10 days and 2 nights, Community Hospitals 12 days and 2 nights, District Hospitals 14 days and 3 nights, and Tertiary Hospitals 28 days and 7 nights, between January and May 2024. Per day/night shift, the enumerators noted whether the whole shift was observed or not. Further details on the data collection operating procedures can be found elsewhere (Nkhoma *et al*., 2025a). In total, the study collected data for 333 HCWs, 677 worker-shift observations, and 42211 individual activities from the sampled facilities.

### Data preprocessing

We implemented a systematic data curation and harmonisation workflow to ensure internal consistency, temporal accuracy, and analytical robustness. (1) Checked and corrected inconsistent or missing date and facility information (e.g., start/end dates, facility closures, duplicate records) to ensure that each record represents a valid observation day at an open facility. (2) Cleaned and harmonised HCW characteristics (e.g., gender, observation day, employment status, highest educational attainment, years in profession, years at the facility) to ensure internal consistency across observation days for the same individual. (3) Corrected abnormal or inconsistent HCW cadre entries and replaced cadre names with standardised definitions aligned with the Health Sector Strategic Plan (HSSP) III (Government of Malawi, 2023). (4) Corrected ambiguous activities (e.g. “Other”, “Nothing”) using informative field notes from data enumerators, resulting in 167 activities labels that include both pre-defined and new frequently observed activities. (5) Corrected implausible activity times and outliers. We investigated long activities, including those exceeding shift lengths, over 3 hours, and in the top 1% of remaining observations, and corrected implausible durations, which were primarily caused by AM/PM errors, overlapping or inherited start/end times, and incorrect end dates. We also removed activities of HCW “not in yet”. (6) Improved consistency of patient information, making sure the binary new patient indicator and the patient number match: i.e., per worker-shift, every unique patient number corresponds to a new patient seen by the worker, and every new patient seen has a unique patient number. (7) Corrected the start or end time of administrative tasks, breaks and other activities to remove the abnormal time overlaps with direct patient care activities.

We found notable cases of 1-minute gaps between recorded activities (81% of 6059 gap observations) and 0-minute activities (16% of 28499 activity observations). The latter resulted from recording the time to the nearest minute. The former could be partially due to the same reason and partially due to real time gaps between two activities, which are difficult to distinguish, however. To tackle the potential underestimation of activity time due to these cases, we conducted further preprocessing. (8) Adjusted times of two adjacent activities by evenly allocating the 1-minute gap, i.e., 30 seconds added to the end time of the proceeding activity and 30 seconds added to the start time of the succeeding activity. For remaining 0-minute activities that have time gaps with adjacent activities, we (9) increased their durations to 20s by adjusting their end time 20 seconds later or start time 20 seconds earlier, depending on the time gaps with neighbouring activities. The 20 seconds is the expected duration of 0-minute activities under the assumption that the real start time and end time are uniformly distributed within recorded start/end time ± 1 minute, and end time is no earlier than start time. The minority cases of gaps longer than 1 minute were checked and treated as Unrecorded time gaps between activities, and nonzero-minute activities were not adjusted because their recorded duration equals the expected duration under the same assumption. See Appendix A.1.4 for more details.

To facilitate stratified analyses across clinically meaningful subgroups, we additionally created categorical variables for cadres, activities, and service areas, referring to the literature (Government of Malawi, 2023) or consulting clinical experts. For the original HCW cadres, the categories include Clinical (including Mental health clinical officer), Nursing and Midwifery (including Psychiatric Nursing Officer), Laboratory, Pharmacy, Health Surveillance Assistant (HSA), Dental, HIV Diagnostic Assistant (HDA), Radiography, and Support Staff (i.e., Hospital Attendant). The mental health staff are categorised into Clinical and Nursing and Midwifery cadres due to anonymity concerns relating to there being only one HCW observed.

For activities, the original labels were first grouped into activity sub-categories then grouped into categories of Patient-facing (activities related to direct patient care, including diagnostic tests by Laboratory staff and drugs dispensing by Pharmacy staff), Administrative, Break, Sleep, HCW training (activities related to HCWs training interns/students/others), Travel (activities related to travel to work, such as HSAs going to communities), Unallocated (activities labelled as “No activity”, meaning time periods not related to any type of work or breaks due to the absence of patients, HCWs temporarily unavailable, power outages, among other reasons), and Unknown (activities with missing information).

For the original service areas, the categories are Emergency care (including Emergency / Intensive care), Inpatient care (including Inpatient – General, Inpatient - Maternity/Gynaecology, and Surgery), and Outpatient care (including Outpatient – General, Outpatient - Maternity/Gynaecology, CD (communicable disease) clinic, NCD (non-communicable disease)/Other clinic, Laboratory, and Pharmacy). Full lists of cadres, activities, and service areas as collected in the TMS study, alongside the categorisation, can be found in Appendix A.1.3.

We also created shift types – Day and Other – to distinguish the time use patterns during different shifts. The Day shift is defined as the observed HCW starting to work between 7am-5pm and the observation day did not cross two dates. All remaining cases were categorised as Other shift, including 24-hour and on-call shifts that could not be clearly differentiated.

#### Preparing the main sample

We prepare the sample for main analyses to include only data of Day shifts that were completely observed, considering that Day shifts have a much larger sample size than Other shifts and partial day observations produce incomplete information for a whole shift. HSAs and Support staff are further excluded as they have less than 10 worker-day observations. The resulting sample includes 227 HCWs and 423 worker-day observations, with at least 10 worker-day observations for each remaining cadre and each worker being observed for a median of 3 days (IQR: 1-3).

Table 1 summarises the characteristics of the HCWs and work-day observations in the main sample. The percentages across the categories per characteristic are consistent between HCWs and worker-day observations, indicating that no subgroup of workers was disproportionately observed in the dataset. Besides, the sampled HCWs have a median of 6-year (IQR: 3-12) professional experience and a median of 4-year (IQR: 1-6) working time at the facility. Most HCWs did not move across multiple service area categories during a day shift (Table 2). Note in public and CHAM health sectors in Malawi, the health workforce distribution among cadres is: Clinical 16%, Nursing and Midwifery 33%, Laboratory 3%, Pharmacy 2%, Dental 0.6%, Mental 0.2%, Radiography 0.8%, Nutrition 0.3%, HSA 44% (She *et al*., 2024; Berman *et al*., 2022; Government of Malawi, 2023). The sampled HCWs overall represent the total health workforce, despite certain discrepancies due to the sampling strategies that have oversampled doctors and specialists, undersampled HSAs, and selected facilities evenly across ownerships, rural and urban areas, and facility types.

**Table 1.**
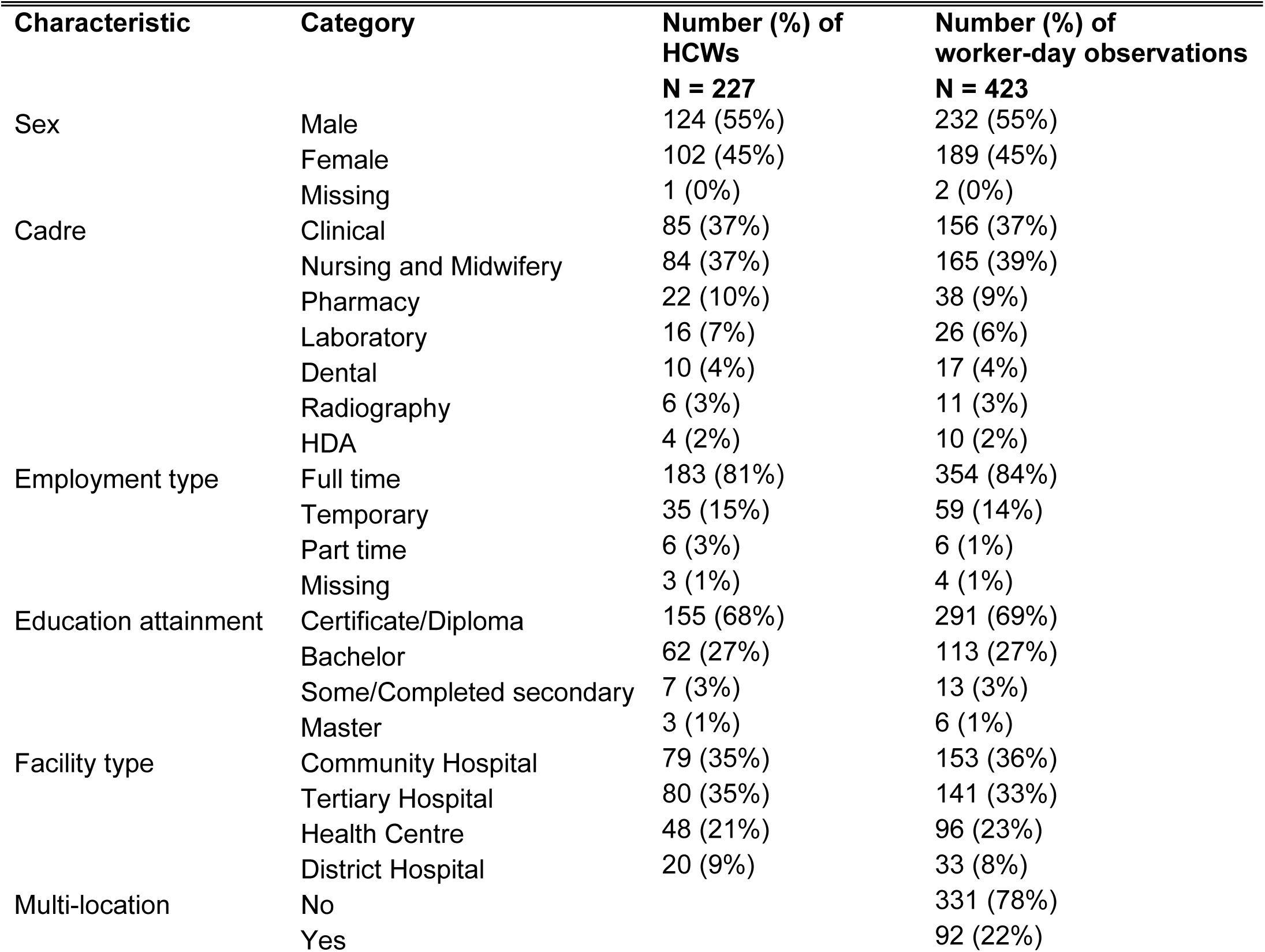
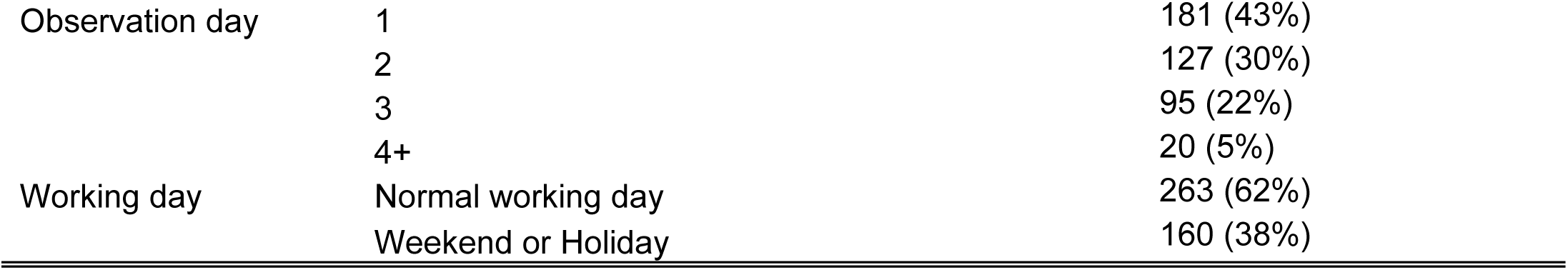
Characteristics of HCW respondents and worker-day observations.

**Table 2.**
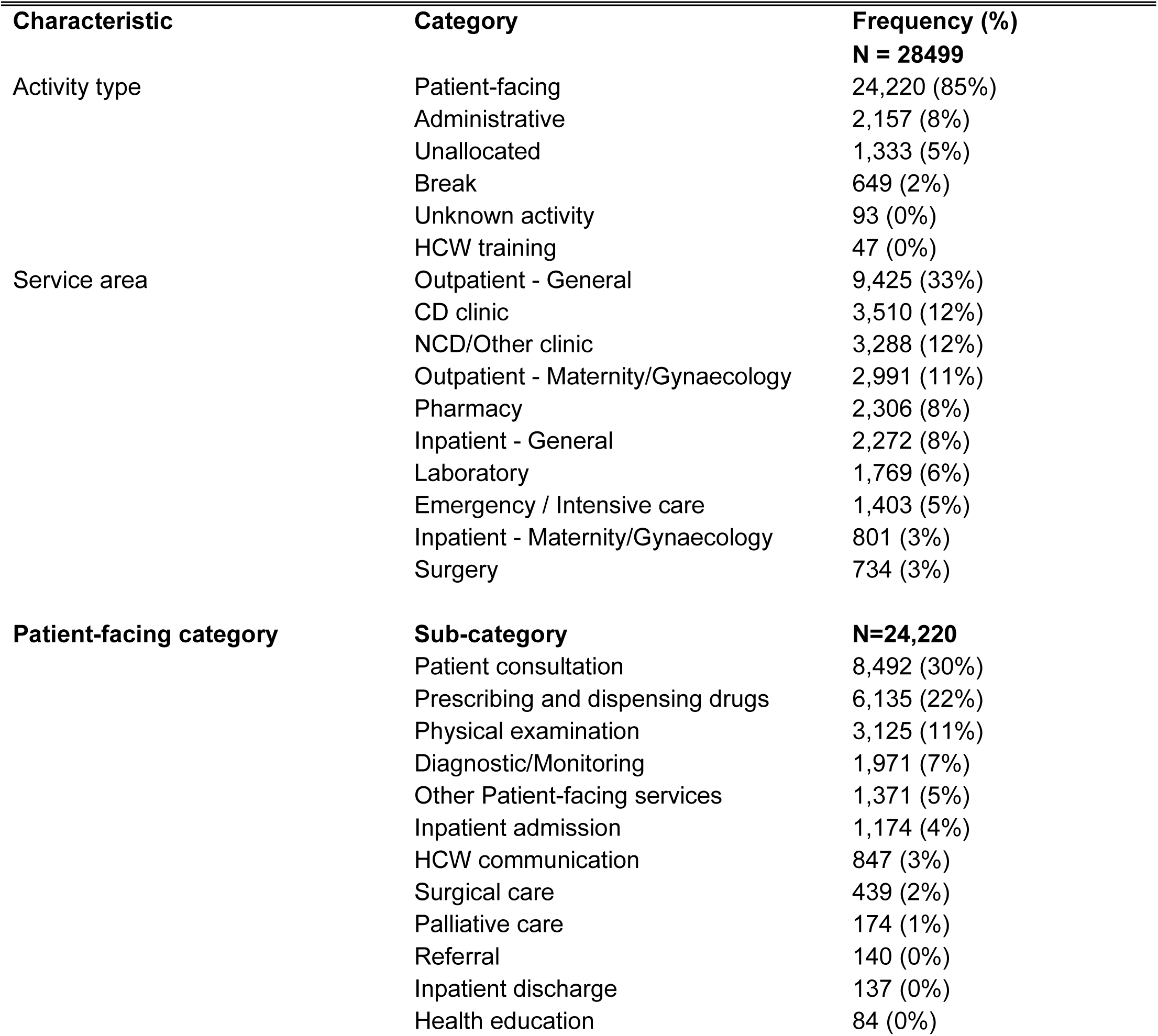
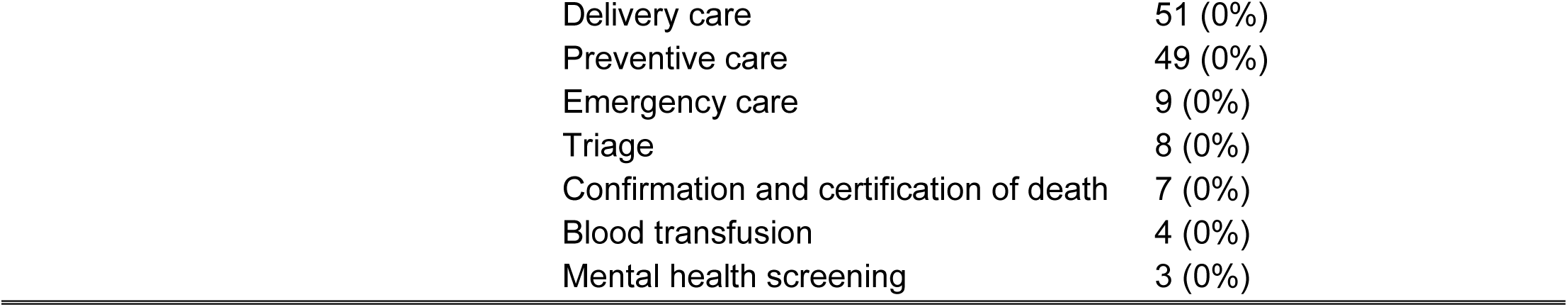
Characteristics of HCW activities.

| Characteristic | Category | Frequency (%) |
| --- | --- | --- |
|  |  | **N = 28499** |
| Activity type | Patient-facing | 24,220 (85%) |
|  | Administrative | 2,157 (8%) |
|  | Unallocated | 1,333 (5%) |
|  | Break | 649 (2%) |
|  | Unknown activity | 93 (0%) |
|  | HCW training | 47 (0%) |
| Service area | Outpatient - General | 9,425 (33%) |
|  | CD clinic | 3,510 (12%) |
|  | NCD/Other clinic | 3,288 (12%) |
|  | Outpatient - Maternity/Gynaecology | 2,991 (11%) |
|  | Pharmacy | 2,306 (8%) |
|  | Inpatient - General | 2,272 (8%) |
|  | Laboratory | 1,769 (6%) |
|  | Emergency / Intensive care | 1,403 (5%) |
|  | Inpatient - Maternity/Gynaecology | 801 (3%) |
|  | Surgery | 734 (3%) |
| Patient-facing category | **Sub-category** | **N=24,220** |
|  | Patient consultation | 8,492 (30%) |
|  | Prescribing and dispensing drugs | 6,135 (22%) |
|  | Physical examination | 3,125 (11%) |
|  | Diagnostic/Monitoring | 1,971 (7%) |
|  | Other Patient-facing services | 1,371 (5%) |
|  | Inpatient admission | 1,174 (4%) |
|  | HCW communication | 847 (3%) |
|  | Surgical care | 439 (2%) |
|  | Palliative care | 174 (1%) |
|  | Referral | 140 (0%) |
|  | Inpatient discharge | 137 (0%) |
|  | Health education | 84 (0%) |

|  |  |
| --- | --- |
| Delivery care | 51 (0%) |
| Preventive care | 49 (0%) |
| Emergency care | 9 (0%) |
| Triage | 8 (0%) |
| Confirmation and certification of death | 7 (0%) |
| Blood transfusion | 4 (0%) |
| Mental health screening | 3 (0%) |

Table 2 summarises the characteristics of the health workers’ activities as observed in the main sample, showing that most activities observed are about Patient-facing services (as listed in the Patient-facing category) across outpatient, inpatient, and emergency care settings.

### Analysis

We generated descriptive statistics to describe HCW time-use patterns on a per-worker, per-day shift basis. Specifically, we quantified: (i) total working time, defined as the elapsed time from an HCW’s arrival at the health facility to log-off, including Break and Unallocated time; (ii) the allocation of working time across activity categories; (iii) patient load, i.e., the number of unique patients seen by a HCW, both overall and at each service area; and (iv) time per patient, i.e., the total time spent on a patient by a HCW (including repeated visits of the same patient to the same HCW during that day shift), both overall and at each service area. For each measure, we provided an overview of the pooled sample and then explore whether patterns vary by HCW cadre, facility type, facility ownership, region, rural/urban area, catchment population size, observation day, workday/weekend or holiday, and service area. The analysis on observation day investigated the Hawthorne effect that may be introduced as the presence of observers may alter the HCWs behaviours, but this bias reduces quickly over time (Bryant and ESSOMBA, 1995; Leonard and Masatu, 2006).

For working time allocation, the total time of each activity category other than Patient-facing was calculated as the sum of all activities in that category, and Patient-facing time was the difference between total working time and the total time of other activity categories, considering that there were no time overlaps within non-patient-facing activities or between Patient-facing and non-patient-facing activities. For time per patient, because time overlaps are expected within Patient-facing activities (e.g. multi-tasking), it was calculated as the sum of non-overlapped, continuous time blocks for a patient. For all measures, statistics include median and interquartile range (IQR 25th percentile-75th percentile).

To calculate the patient load and time per patient, we exclude worker-day observations involving HCWs working in multiple service area categories during a day shift. These observations are not directly comparable to single-service area contexts, as working time distributed across heterogeneous service areas would introduce systematic bias, particularly in estimates of daily patient load. Restricting the analysis to single-service area observations ensures internal consistency in time-patient mapping. The retained sample (78% of observations) exhibits comparable distributions across key characteristics (Appendix Table A.2).

Differences in medians between subgroups were further assessed using pairwise Mood’s median tests (p < 0.05), a non-parametric approach suitable for non-normal distributions.

Finally, sensitivity analyses are performed for working time allocation and time per patient using data prior to preprocessing steps (8) and (9), to investigate the influence of frequent 1-minute gaps and 0-minute activities on time-use estimates. Such artefacts may introduce fragmentation or misallocation of time across activities. Daily working time and patient load are unaffected by these preprocessing steps and are therefore not included in the sensitivity analysis.

## Results

### Overview

Overall, the HCWs worked 7.35 hours (IQR 4.40-8.35) per day shift. Within the total working time, Patient-facing time was 2.82 hours (IQR 1.89-3.97), accounting for 47.52% (IQR 30.44%-66.87%); Administrative time was 0.30 hours (IQR 0.00-1.23), with 5.21% (IQR 0.00%-17.50%); Break time was 1.25 hours (IQR 0.00-2.12), with 17.89% (IQR 0.00%-27.97%); and Unallocated time was 0.72 hours (IQR 0.02-1.92), with 12.00% (IQR 0.21%-28.62%). The unrecorded time gaps were negligible, with 0.00 hour (IQR 0.00-0.19) and 0.00% (IQR 0.00%-3.79%).

By investigating the reasons for Unallocated time, where 65% of activities in this category had such information provided, it is found that the most frequent reason was no patient to attend to or waiting for patients to come and this accounts for 71% of the total Unallocated time of these activities. (Appendix A.2.2).

The median number of patients seen per HCW per day was 21(IQR 12-35) across outpatient care areas, 12 (IQR 7-18) across inpatient care areas, and 14 (IQR 10-23) in emergency care, with the time per patient at 3 minutes (IQR 1.0-6.5), 6 minutes (IQR 2.5-14), and 10 minutes (IQR 5-20), respectively.

### Subgroup analyses

Next, we inspected HCWs working time use patterns by HCW cadres, facility characteristics, service areas, among others.

#### The daily working time

As shown in Figure 1, the daily working time had substantial variations within each cadre, facility type, ownership, region, rural/urban area, high/low catchment population size, normal working day/weekend or holiday, and observation day category, indicated by wide interquartile ranges (4-9 hours) and overall ranges (1-11 hours) for each subgroup. However, the differences between subgroups of each characteristic were overall insignificant. The IQRs mostly overlapped. Furthermore, the median values of most subgroups have no significant difference according to the pairwise Mood’s median tests (Appendix A.2.1). This include the observation day category, indicating that the daily working time is less prone to bias due to the Hawthorne effect. Nonetheless, the tests found that HCWs at Community hospitals worked significantly longer than at Health Centres and Tertiary hospitals (by 1.75 hours and 0.95 hours, respectively), and those in the Central region worked significantly shorter than in the Northern and Southern regions (by 1.28 and 1.55 hours, respectively).

**Figure 1.**
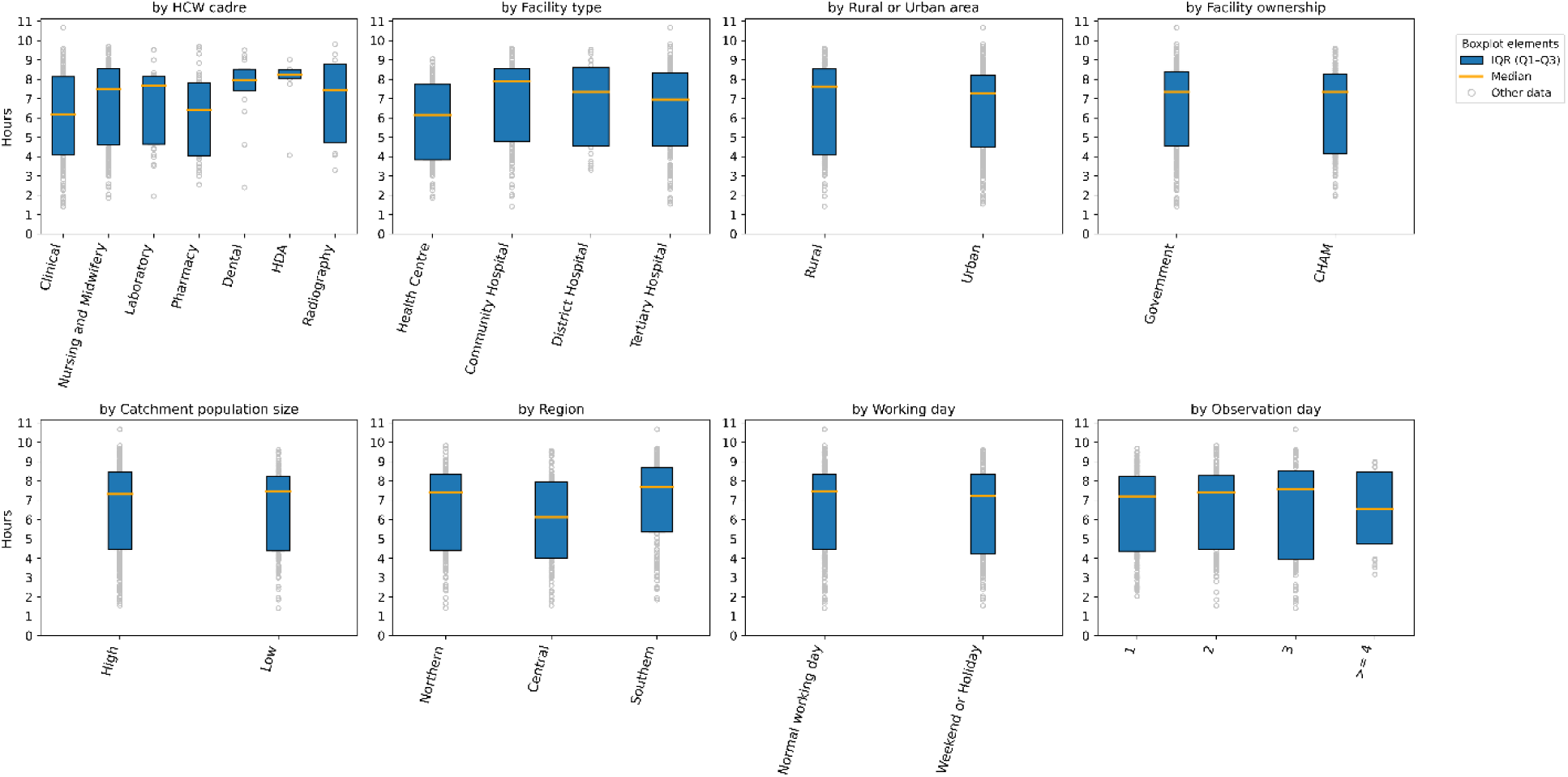
The daily working time (hours).

#### Working time allocation

Figure 2 shows the daily working time allocation across Patient-facing, Administrative, Break, and Unallocated categories, alongside the unrecorded time gaps. Note HCW training and Unknown activity categories are excluded due to minimal recorded time. Across the characteristics and subgroups of each characteristic, it is consistent that Patient-facing time had the highest proportions, with median values ranging from 40% to 60%. The remaining categories had much lower proportions with median values below 25%, where Break and Unallocated proportions were comparable, and Administrative proportions were the lowest in most categories of characteristics. The unrecorded time gap proportions were overall negligible. Although, substantial variations within each subgroup were notified by the wide IQRs and/or overall ranges: e.g., for Patient-facing time proportion of Clinical cadre, the IQR is 40%-79% and the overall range is 6%-99%.

**Figure 2.**
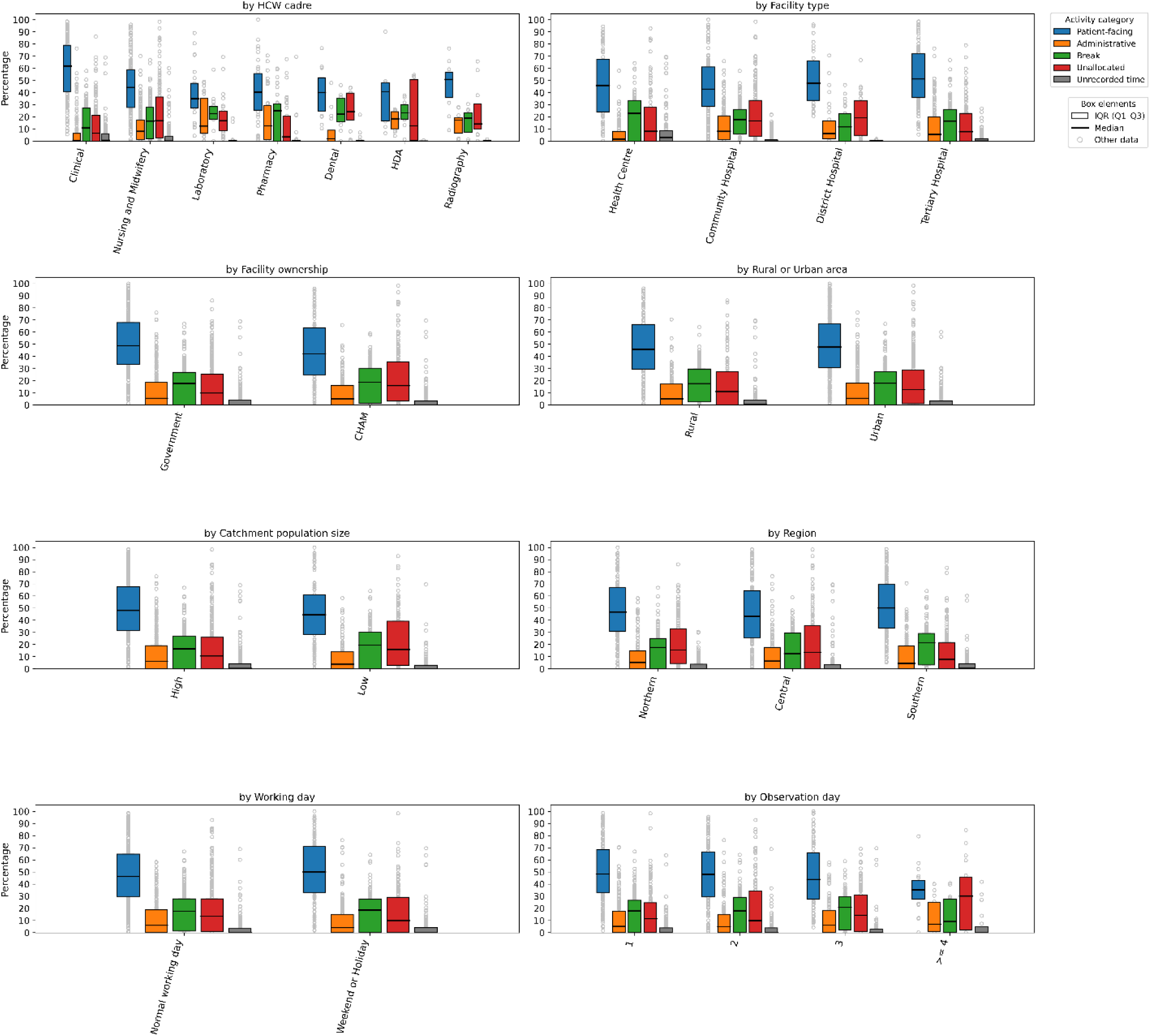
Daily working time allocation (%)

Furthermore, for each activity category except Break, significant differences between subgroups were identified through the pairwise Mood’s median tests (Appendix A.2.1). For Patient-facing time, Clinical cadre had a much higher proportion than Nursing and Midwifery by 18%, Laboratory by 27%, and Pharmacy by 21%; Government facilities had a higher proportion than CHAM facilities by 7%; observation day 1 and day 2 had higher proportions than day 4 and later by 13%, but this difference may be less reliable as the latter had only 20 observations as compared to more than 100 observations for earlier observation days. For Administration time, Clinical cadre had lower proportions than Nursing and Midwifery by 8%, Laboratory and Pharmacy by 12%, and Radiography by 17%, but Radiography cadre had only 11 observations; Health Centres had a lower proportion than Community hospitals by 7%. For Unallocated time, Government facilities had lower proportions than CHAM facilities by 6%, and the Southern region had lower proportions than the Northern region by 7%.

#### The daily patient loads

Daily patient loads per HCW varied widely across all characteristics, as reflected by wide IQRs and overall ranges within each subgroup. While some clinical HCWs saw more than 150 patients per day, others saw fewer than 10. Despite this variability, median patient loads were broadly comparable across most subgroups of each characteristic. Significant differences were primarily observed by service area and facility ownership, with General outpatient clinics having the highest median (29 patients) whereas Maternal/Gynaecological inpatient wards having the lowest median (9 patients), and Government facilities seeing about 5 more patients than CHAM facilities. Further details can be found in Appendix A.2.3.

Here we mainly present the patient loads with the service area fixed as shown in Figure 3, considering the significant differences across service areas. As highlighted by the pairwise Mood’s median tests (Appendix A.2.1), HCWs at General outpatient clinics had significantly different patient loads between categories of characteristics including cadre, facility ownership, region, and normal working day/weekend or holiday. More specifically, Clinical cadre saw 25 more patients per day than Nursing and Midwifery cadre; HCWs at Government facilities saw 19 more patients per day than at CHAM facilities; HCWs in the Northern region saw 29 more patients per day than the Central region; and HCWs working in weekends/holidays saw 12 more patients per day than in normal working days. By contrast, in other service areas, no significant differences were found between the categories of each characteristic.

**Figure 3.**
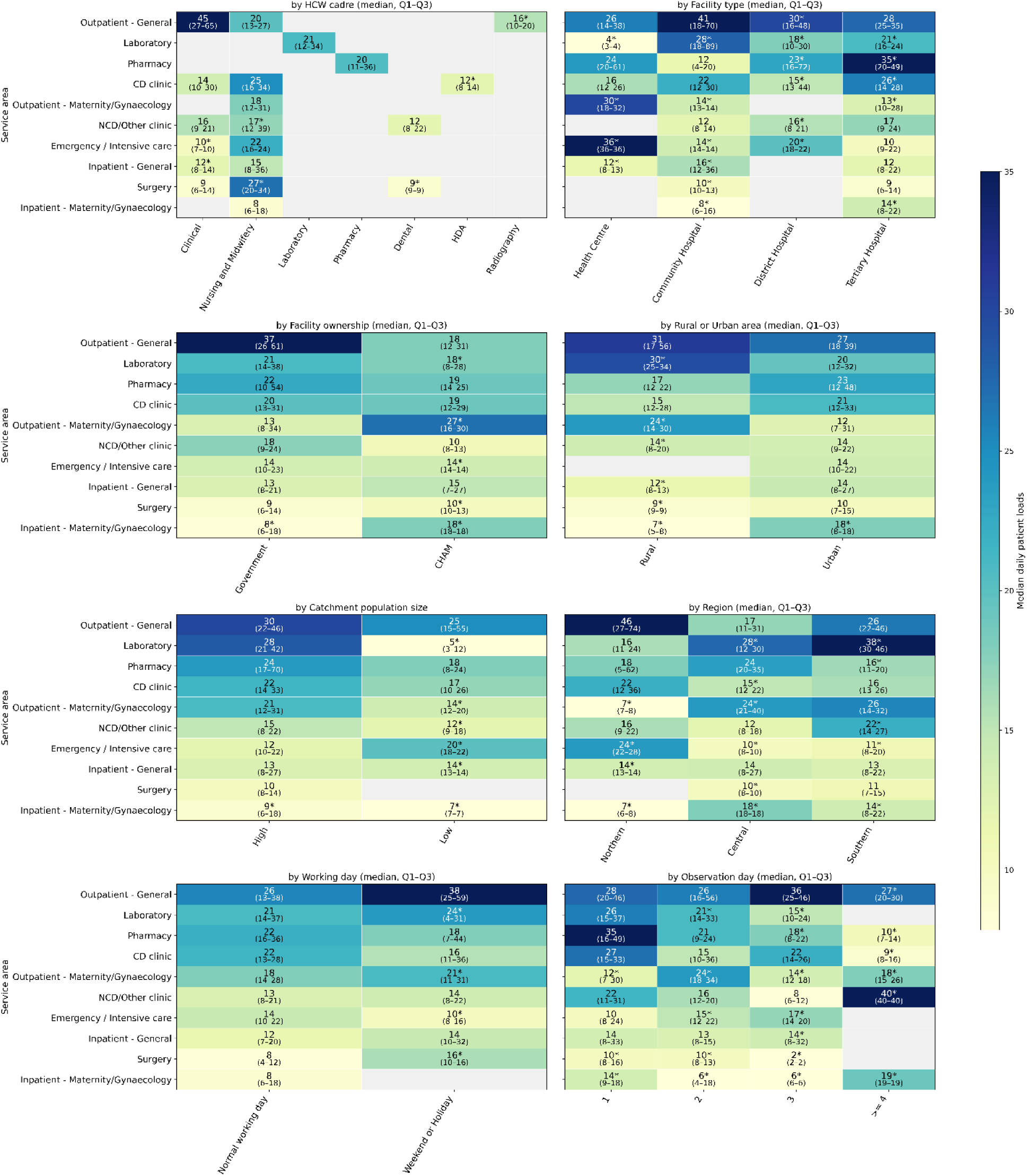
Daily patient loads per service area. Entries with fewer than 10 worker-day observations are annotated with stars.

#### Time per patient

As the same with above results, time per patient varied substantially in subgroups of key characteristics as shown by wide IQRs and overall ranges: short tasks (e.g., sending patients to do malaria tests or drug dispensing) took under 1 minute, whereas some surgeries exceeded 200 minutes. But in contrast to patient load, median time per patient differed significantly across most subgroups and characteristics. Overall, Dental, HDA, and Radiography cadres had the highest time per patient, followed by Nursing and Midwifery, then Clinical and Laboratory cadres, with Pharmacy cadres the lowest. Time per patient was higher in Tertiary hospitals compared to District, Community hospitals, and Health Centres, and higher in CHAM facilities than in Government facilities. Urban settings showed higher values than rural areas, while regionally the Northern region had the lowest time per patient, followed by Central, with Southern the highest. By service area, the highest time per patient was observed in Surgery clinics, Emergency care units, and Maternal/Gynaecological inpatient wards, followed by CD clinics, Maternal/Gynaecological outpatient clinics, NCD/Other clinics, and General inpatient wards, while Laboratories, General outpatient clinics, and Pharmacies had the lowest. See full details in Appendix A.2.4.

Figure 4 further presents time per patient across characteristics with the service area held constant. While patterns for General outpatient clinics broadly mirror the overall trends as above, distinct disparities emerge across other areas. Clinical staff consistently recorded higher time per patient than Nursing and Midwifery staff in NCD/Other clinics, General inpatient wards, Emergency care units, and Surgery clinics. Community hospitals showed higher time per patient in Pharmacies, Maternal/Gynaecological outpatient clinics, General inpatient wards, and Maternal/Gynaecological inpatient wards. Government facilities exceeded CHAM facilities in Maternal/Gynaecological outpatient clinics. Rural areas exhibited higher values in Pharmacies, NCD/Other clinics, and both General inpatient and Maternal/Gynaecological inpatient wards. Regional differences are also evident, with the Southern region generally showing lower values in Maternal/Gynaecological and General inpatient wards, while the Northern region showed higher values in Maternal/Gynaecological outpatient clinics.

**Figure 4.**
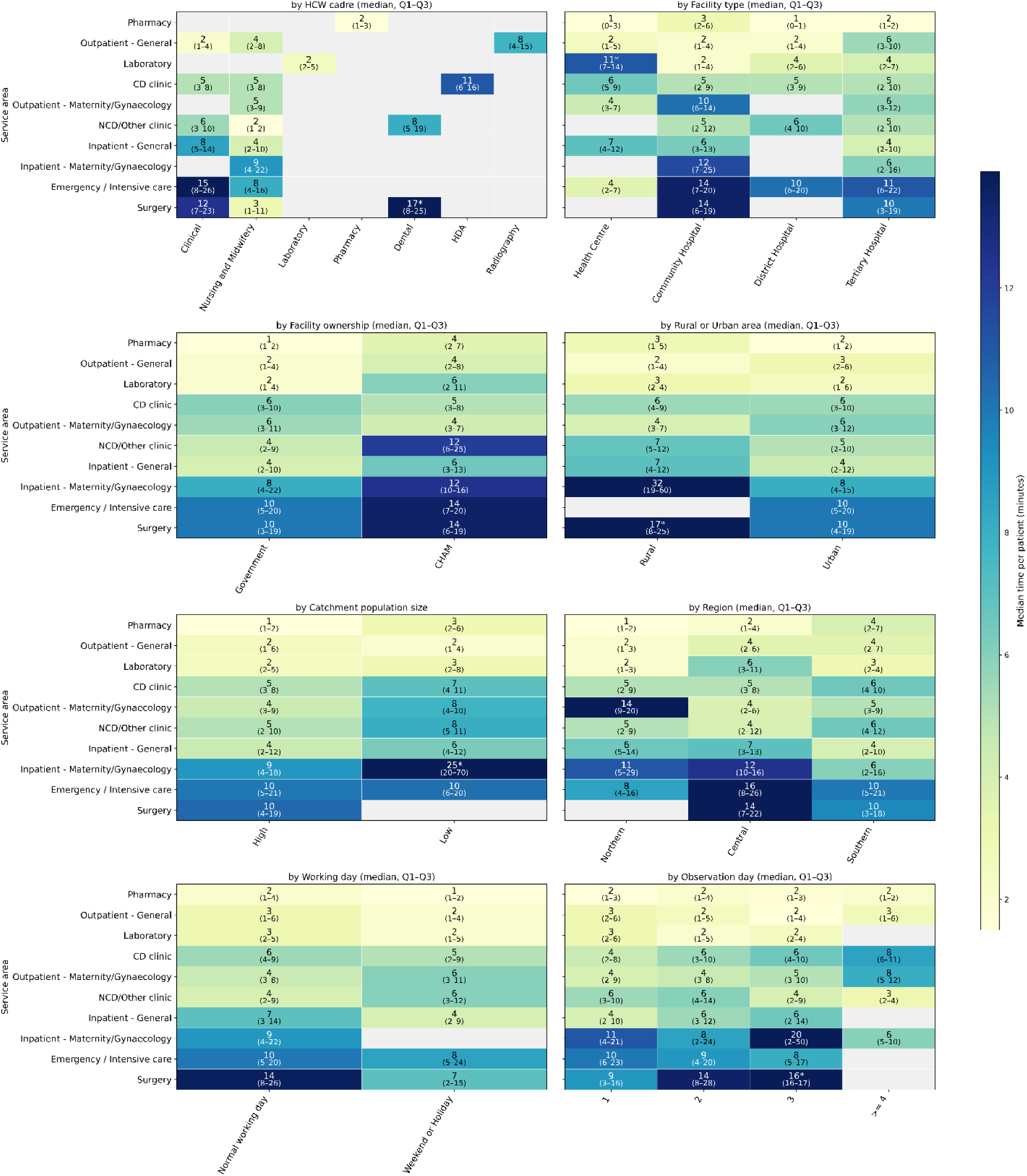
Time per patient per service area. Entries with fewer than 10 patient observations were annotated with stars.

Additionally, facilities serving smaller catchment populations had higher time per patient in CD, NCD/Other, Maternal/Gynaecological outpatient clinics, and Pharmacies. Time per patient was Generally higher on normal working days than weekends or holidays for General outpatient clinics, Pharmacies, General inpatient wards, and Surgery clinics, whereas the opposite pattern is observed in NCD clinics.

### Sensitivity analysis

Without data preprocessing on time gaps and 0-minute activities, the overall unrecorded time gaps account for 3.48% (IQR 1.23%-8.00%) of total daily working time, compared to 0% (IQR 0%-3.79%) in the main analysis. This corresponds to a slightly lower Patient-facing proportion that is 43.53% (IQR 28.48%-61.67%), compared to 47.52% (IQR 30.44%-66.87%) in the main analysis. Accordingly, the time per patient across characteristics had very minor, non-significant decreases; however, disparities are consistent with the main findings. Further details can be found in Appendix A.3.

## Discussion

Existing time-and-motion studies on HCW time use in Malawi and other low- and middle-income countries have typically been limited in scope, focusing on single cadres (e.g., community health workers, doctors, or nurses), a limited number of facilities, or specific diseases and programmes at community or primary care levels (Muho *et al*., 2025; Aron *et al*., 2023; Chinkhumba *et al*., 2022; Muho, Peshkatari and Wyss, 2022; Brar *et al*., 2021; Chebolu-Subramanian *et al*., 2019; Singh *et al*., 2018; Bonfim *et al*., 2016; Tilahun *et al*., 2017; Gardner *et al*., 2010; Gottschalk and Flocke, 2005; Pizziferri *et al*., 2005; Sessions *et al*., 2019; Jafry *et al*., 2016). This limitation has constrained system-level understanding of workforce utilisation in resource-limited settings. Addressing this gap, our study provides, to our knowledge, the first comprehensive, system-wide, multi-cadre evidence on HCW time use across service areas, facility types, and geographical locations (rural/urban areas, regions) in Malawi, offering a more generalisable and policy-relevant evidence base for LMIC health systems.

Overall, median daily working time was 7.35 hours (including breaks), consistent with previous estimates for HCWs (Berman *et al*., 2022; Government of Malawi, 2018). However, it falls short of the contracted schedule by approximately 1.65 hours. The working hours also differed significantly across facility types and regions, with HCWs at Community hospitals working longer than those at Health Centers and Tertiary Hospitals, and those in the Central region working shorter than those in the Northern and Southern regions. The HCWs spent most of their time on direct patient care (48% of daily working time), with Break time consistent with the contracted duration (18%) and a minor administrative burden (5%). But there was Unallocated time on a similar magnitude of Break time, which is largely due to no patient to attend to or waiting patients to arrive according to informative field notes. There are also variations across facility ownership and region, as Government facilities and the Southern region have lower Unallocated time proportions than CHAM facilities and the Northern region, respectively.

Daily patient loads in outpatient care – a median value of 21 patients per HCW – were relatively high compared to other Sub-Saharan Africa countries, which is consistent with findings from patient load studies (Daniels *et al*., 2025; Lilford *et al*., 2025). Furthermore, patient loads varied across outpatient, inpatient, and emergency care settings, where outpatient settings saw more patients than the others. This is consistent with the national Health Management Information System (HMIS) data from Malawi, which shows that outpatient visits far exceed hospital discharges (She *et al*., 2024). Interestingly, our results do not suggest significantly different per-HCW patient loads across facilities at primary, secondary, and tertiary levels of care either overall or in specific service areas. This may contrast with the literature that emphasises hospital centrism and bypass of primary care, leading to overcrowded higher-level hospitals and under-utilised lower-level facilities (Li, Chen and Khan, 2021). However, it is consistent with a recent study on 10 LMIC cities suggesting that staffing levels generally scale with total patient volume, such that per-HCW caseloads converge across facility types (Lilford *et al*., 2025). It is also notable that General outpatient clinics saw more patients on weekends/holidays than on normal working days despite comparable working time and patient-facing time; this may reflect differences in patient demand, as reduced work and caregiving constraints lower the opportunity cost of seeking care (Ensor and Cooper, 2004), or may be due to fewer HCWs working on weekends/holidays, which requires further investigation on patient care seeking behaviour and staffing levels.

Meanwhile, the overall time per patient was relatively low across service areas, although inpatient wards, emergency care units, and surgical clinics with median 9 to 11 minutes had a higher time per patient than outpatient areas with median 2 to 6 minutes. In particular, the median 2 to 4 minutes of Clinical/Nursing and Midwifery time in General outpatient clinics – was among the lowest consultation time findings for SSA and international countries (Daniels *et al*., 2025; Irving *et al*., 2017). The time per patient across service areas was also much shorter than the standard time requirements according to Malawian government and WHO documents. For instance, an outpatient consultation would need more than 10 minutes of Clinical/Nursing and Midwifery time, and inpatient ongoing monitoring and emergency care would need more than 30 minutes of their time (Berman *et al*., 2022; Government of Malawi, 2023; Mziray, Gorgens and McCauley, 2017). However, these discrepancies should be carefully interpreted. First, our results exhibited substantial variations in each subgroup of characteristics at each service area, indicating the various nature of services and patient health conditions. Second, the time per patient here is calculated from every single worker-day observation, rather than from the total time that a patient may be attended to by multiple HCWs at multiple locations during one visit to the facility, where the latter would be higher.

Our time per patient results also show important disparities regarding facility type, facility ownership, and geographical characteristics. Overall and in General outpatient settings, Central hospitals, CHAM facilities, urban settings, and the Southern region had higher time per patient, where the finding on CHAM facilities is consistent with the literature (Tafesse *et al*., 2025). However, these disparities patterns are not consistent across other settings. Contrastingly, Community hospitals showed higher time per patient in General inpatient care, Maternal/Gynaecological outpatient and inpatient care, and Pharmacies; Government facilities had higher time per patient in Maternal/Gynaecological outpatient care; Rural areas exhibited higher time per patient in Pharmacies, NCD/Other clinics, and Maternal/Gynaecological inpatient care. Regional differences are also service-specific: the Southern region showed lower time per patient in both General and Maternal/Gynaecological inpatient care, while the Northern region exhibited higher time per patient in Maternal/Gynaecological outpatient care. Complementing existing literature (Misu, Gasbarro and Alam, 2025; Kruk *et al*., 2018; Campbell *et al*., 2013; Zere *et al*., 2007), these findings demonstrate that health inequity issues across health system levels and geographical locations in low-income countries can be service-specific, for which interventions to improve health equity should be carefully designed and evaluated.

It is important to note that these HCW time-use patterns are based on median estimates; however, the wide IQRs and overall ranges within subgroups indicate substantial heterogeneity, reflecting the diverse nature of health service provision and patient conditions.

The observed unallocated time and shortfall relative to contracted working hours suggest under-utilised capacity within the current health workforce, although the extent varies by facility type, ownership, and region. As this time likely reflects periods without patients, and given the combination of high patient loads and low time per patient, there is scope to increase service time per patient if demand were more evenly distributed throughout the day - for example, through improved appointment organisation. This may, in turn, contribute to improved process quality and patient-perceived quality of care (Wang *et al*., 2022; Kim *et al*., 2021; Kruk *et al*., 2018). Nonetheless, sustained improvements will require greater investment in human resources for health and expansion of the workforce to meet growing demand. There should also be simultaneous efforts to improve availabilities of medical consumables, equipment and infrastructure that limit the effective capacity of HCWs (Klootwijk *et al*., 2025; Mohan *et al*., 2024; Mueller *et al*., 2011), and to facilitate balanced healthcare seeking reducing barriers such as direct travel time and cost and opportunity cost due to time away from income-generating activities (Dawkins *et al*., 2021; Abiiro, Mbera and De Allegri, 2014).

To ensure robust and analytically consistent results, rigorous data curation and preprocessing was implemented, including refinement of broad activity categories (e.g., “Other”) using field notes, standardisation of HCW cadres, service locations, and activity classifications, and systematic treatment of time overlaps and unrecorded gaps. This process provides practical methodological insights for future time and motion study design, particularly the value of clearer and more granular activity definitions and higher-resolution time recording (e.g., inclusion of seconds) to better capture short-duration tasks. The results of our sensitivity analyses further strengthen confidence in our findings by showing that our data preprocessing decisions do not materially influence our results.

This study has a few limitations. First, as an observational time-and-motion study, there is a potential for behaviour modification among HCWs (i.e., the Hawthorne effect), which could bias observed time-use patterns. However, analyses across observation days revealed no significant or systematic differences, suggesting that any such effect is limited in this study. Second, observing only present HCWs may bring selection bias to the estimates if absent HCWs’ behaviours differ systematically (Scheffler *et al*., 2016); further investigation on the impact of absenteeism is required. Third, this study used categorised HCW cadres and service areas for analytical clarity, but more granular classifications were also included in the data. Analyses of finer groups would provide more detailed information and/or may reveal hidden disparities due to coarser categorisation (in particular for the time per patient results). Such analyses can be simply reproduced by focusing on finer groups of characteristics but should be carefully interpreted considering the limited sample size. See examples of Clinical and Nurse and Midwifery staff at General outpatient clinics in Appendix A.2.5. Finally, most of the data were collected during the hot and rainy season (typically November to April) in Malawi; therefore, the findings may be less applicable for dry season, when patient health care seeking behaviours, health care demand, and HCW time use patterns may differ (Tani *et al*., 2016; Ewing *et al*., 2011). Within the study period (January to May), no clear temporal differences were observed (Appendix A.2.6), although the study was not specifically designed to detect seasonal effects.

Future research should extend beyond descriptive analyses by incorporating a broader set of potential determinants - such as staffing density at each facility, task shifting, shift type (e.g., night, on-call, 24-hour shift), HCW characteristic (e.g., years of experience, education attainment, contract type), and importantly, patient health condition - and applying multivariable regression and Structural Equation Modelling to jointly account for these factors and better explain observed disparities in HCW time use. However, these techniques would require careful interpretation of the association or causality (Lewer *et al*., 2025). Where feasible, longitudinal data would further strengthen such analyses and support more robust policy recommendations.

## Conclusion

Based on a recent TMS on Malawian HCW time use, this exploratory analysis found that overall HCW working time was lower than contracted hours. HCWs spent most of their time on direct patient care, with a minor administrative burden and breaks that align with the contract, but also unallocated time comparable to break time. Daily patient loads per HCW were high, with relatively low time spent per patient, and substantial variation was observed across cadres, facility and geographic characteristics, and service areas. Overall, we presented a holistic picture of HCW time use in Malawi, contributing to the evidence base in LMIC settings and informing health systems planning and modelling. The gap between contracted and actual working time, alongside observed unallocated time, indicates potential to improve utilisation of limited HCW capacity: through better organisation of patient appointments to increase time per patient and overall service delivery. However, sustained improvements require increased investment in human resources for health and workforce expansion to meet growing demand. Future research should build on these findings by incorporating additional determinants (e.g., staffing, HCW characteristics, shift types, and patient case-mix) and applying multivariable and structural modelling approaches, with longitudinal data, to more rigorously explain observed disparities and strengthen policy-relevant inference.

## Appendix

### A.1 Auxiliary methods

#### A.1.1 The sampled facilities

Here we present the characteristics of the sampled facilities in Table A.1.

**Table A.1.** Characteristics of sample facilities.

| Variable | Measure | Number (%) of facilities<br>N = 27 |
| --- | --- | --- |
| Facility location | Rural | 10 (37.04%) |
|  | Urban | 17 (62.96%) |
| Region | Central | 8 (29.63%) |
|  | Northern | 9 (33.33%) |
|  | Southern | 10 (37.04%) |
| Facility ownership | CHAM | 11 (40.74%) |
|  | Government | 16 (59.26%) |
| Facility type | Health Centre | 9 (33.33%) |
|  | Community Hospital | 10 (37.04%) |
|  | District Hospital | 3 (11.11%) |
|  | Tertiary Hospital | 5 (18.52%) |
| Catchment population* | High | 17 (62.96%) |
|  | Low | 10 (37.04%) |

\* Catchment population is an estimated population assigned to each health facility, representing the number of people expected to be served by that facility. Here facilities were classified as “High” if their catchment population exceeds the sample mean, and “Low” otherwise.

Although we have collected data on 30 facilities from the field, we only use data from 27 facilities for the main analysis due to inconsistent information between sampling and data collection. As one selected facility (Makwasa Estate Clinics, a private health centre) is temporarily closed during the study, an additional facility (Mianga Estate Clinic, another private health centre) is included in the data collection because the patients are diverted there, resulting in 30 facilities in total, rather than 29 facilities as sampled. Due to incorrect ownership information in the master facility list, these two facilities are found to be private instead of governmental. Besides, one health centre (Mlolo Dispensary) is found to be a health post during the data collection, which has only health surveillance assistant staff. Therefore, these 3 facilities are excluded from the main sample.

Please also note that during sampling stratification, region is not considered. Southern region has 3 central hospitals (Queen Elizabeth Central Hospital, Zomba Central Hospital, Zomba Mental Hopsital) selected, resulting in more observations of Clinical and Nursing cadres in that region.

#### A.1.2 The main sample excluding multiple-location worker-day observations

Following the main sample characteristics summarised in Table 1, here we provide the description of the main sample that excludes multiple-location worker-day observations for the calculation of daily patient loads and time per patient at each service area. This filtered sample accounts for 84% of HCW respondents in the main sample and 78% of the worker-day observations, with a median of 3 days (IQR: 2-3) observed for each worker. The distributions across categories of characteristics are consistent with the main sample. The presented facilities are also the same as the main sample.

**Table A.2.**
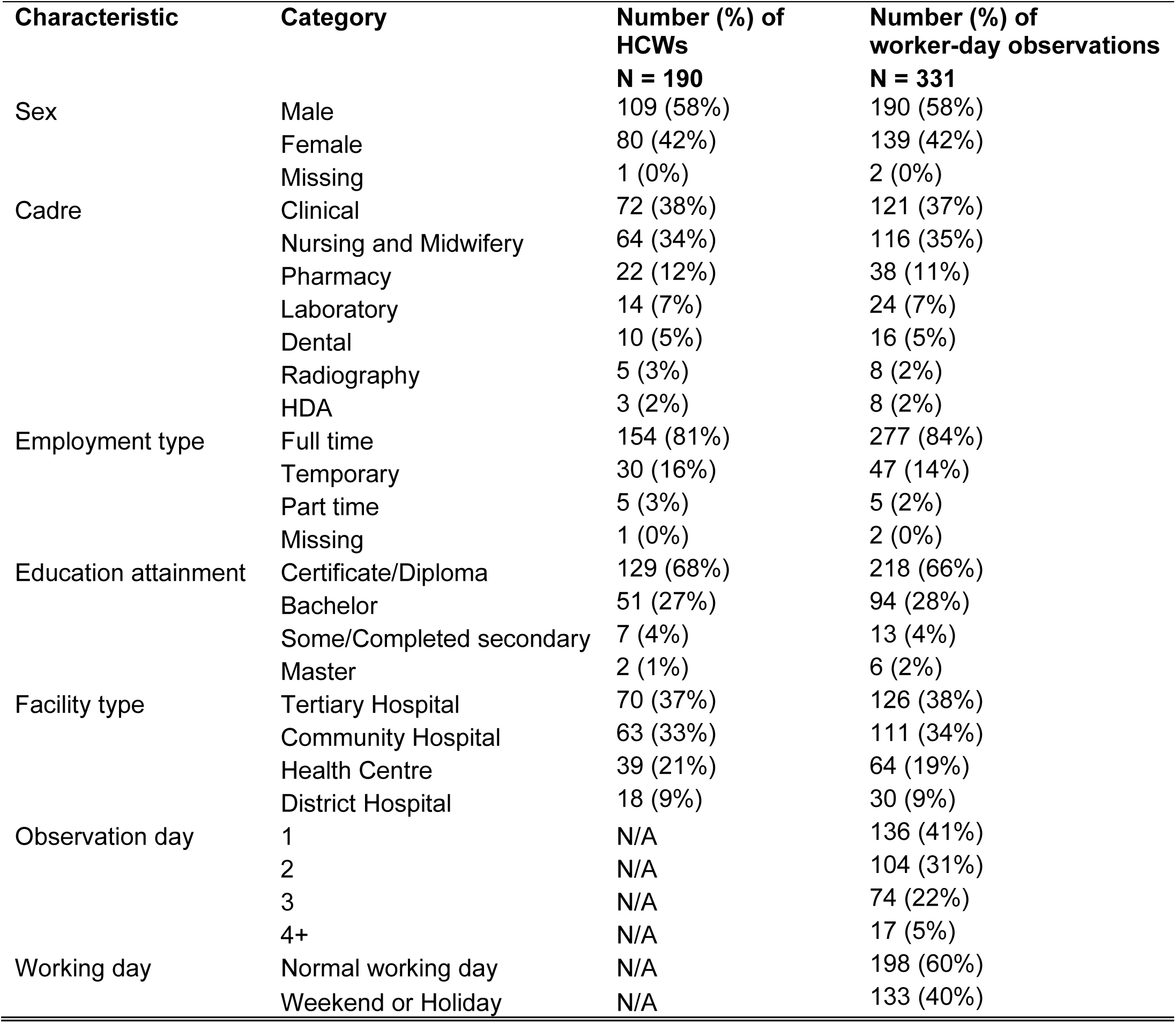
Characteristics of HCW respondents and worker-day observations.

#### A.1.3 Important variables categorisation

The full lists of HCW cadres, activities and clinics/wards/departments as collected in the time-and-motion study, and the categorisation for each can be found in the file “Cadre, Clinic, and Activity categorisation.xlsx” in this repository: 10.5281/zenodo.19852416.

#### A.1.4 Time gaps and 0-minute activities

Before the preprocessing steps (8) and (9), there were 6056 observations of time gaps (i.e., the positive time difference between the start time of each task and the latest end time of all preceding tasks) in the main sample, of which 81% were 1-minute gaps and 19% were gaps greater than 1 minutes. The two preprocessing steps had dealt with all 1-minute gaps. Correspondingly, Figure A.1 below shows the activity durations portions before and after the preprocessing. The remaining 0-minute activities are not adjusted because they have no time gaps with neighbouring activities, meaning their time was fully covered by other activities. These cases can be ignored because of very limited impact on the results as expected.

**Figure A.1.**
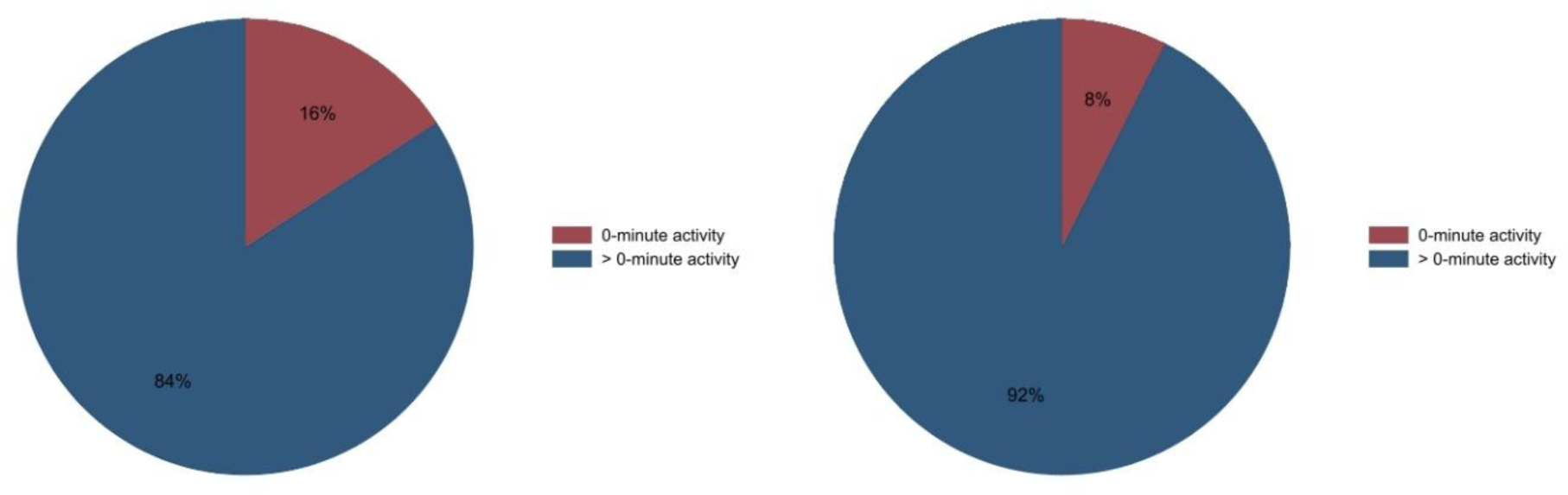
Activity durations proportions before and after preprocessing (N=28499).

### A.2 Auxiliary results

#### A.2.1 Results of pairwise Mood’s median tests

Given the great volume of results from pairwise Mood’s median tests, please find all test results (for daily working time, working time allocation, daily patient loads, and time per patient) in this public repository: https://doi.org/10.5281/zenodo.19852416. Note if the median difference between two subgroups has passed the test at p < 0.05 (i.e., the null hypothesis of equal median values between two subgroups is rejected), it will be treated as significant difference.

#### A.2.2 Details of Unallocated time

By investigating the field notes for activities categorised as “Unallocated” (i.e., those labelled as “No activity” in the data), we found that, among the Unallocated observations with informative field notes, 71% of Unallocated time was due to no patient in presence (See details in Figure A.2 below).

**Figure A.2.**
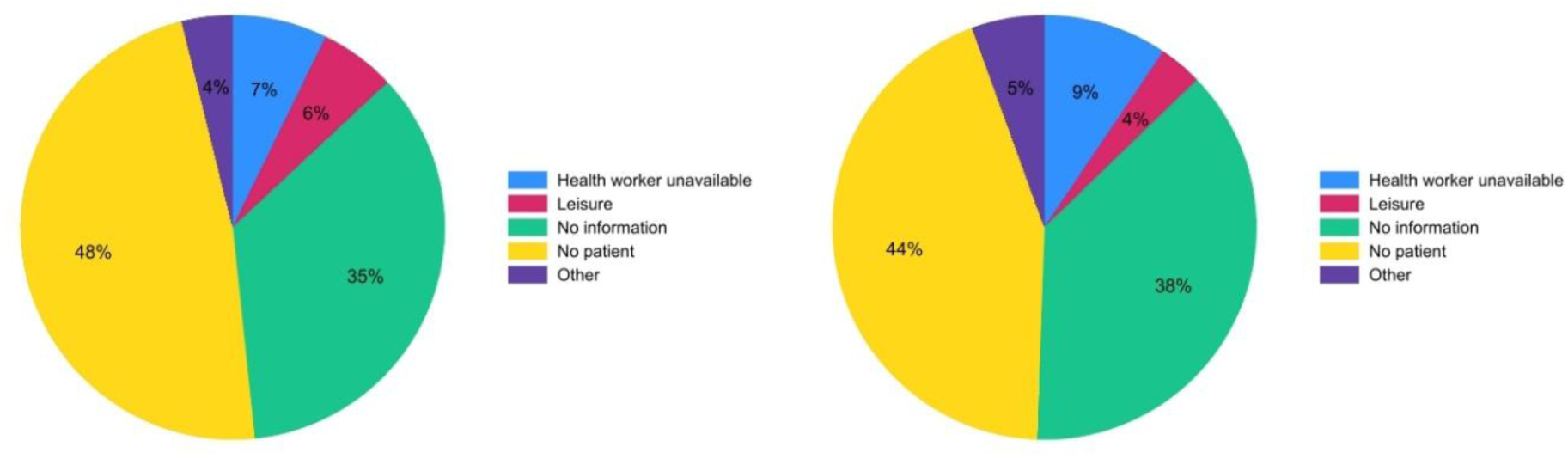
Distribution of Unallocated frequencies and time (N=1,333). The frequency and time proportions of informative observations (excluding “No information”) are 64.7% and 62.3%.

#### A.2.3 Daily patient loads: univariate analysis

Figure A.3 describes the number of patients seen per worker per day across characteristics and categories of each characteristic. As indicated by the wide IQRs and overall ranges, the daily patient loads varied substantially within each subgroup of cadres, facility types, ownerships, regions, rural/urban area, high/low catchment population size, normal working day/weekend or holiday, observation day categories, and service areas. A Clinical HCW was able to see more than 150 patients per day in the General outpatient clinic of a Community Hospital, whereas some others across different characteristics had seen less than 10 patients per day. However, the median patient loads are comparable across the categories of most characteristics. As found through pairwise Mood’s median tests, significant differences mainly took place at different service areas and between Government and CHAM facilities. HCWs at General outpatient clinics had seen most patients with a median value of 29 patients, significantly higher than in NCD/Other clinics, Emergency care units, and Surgery clinics; meanwhile, please note some NCD/other clinics are within General outpatient clinics. HCWs at Government facilities had seen significantly more patients than in CHAM facilities, with a median difference of 5 patients.

**Figure A.3.**
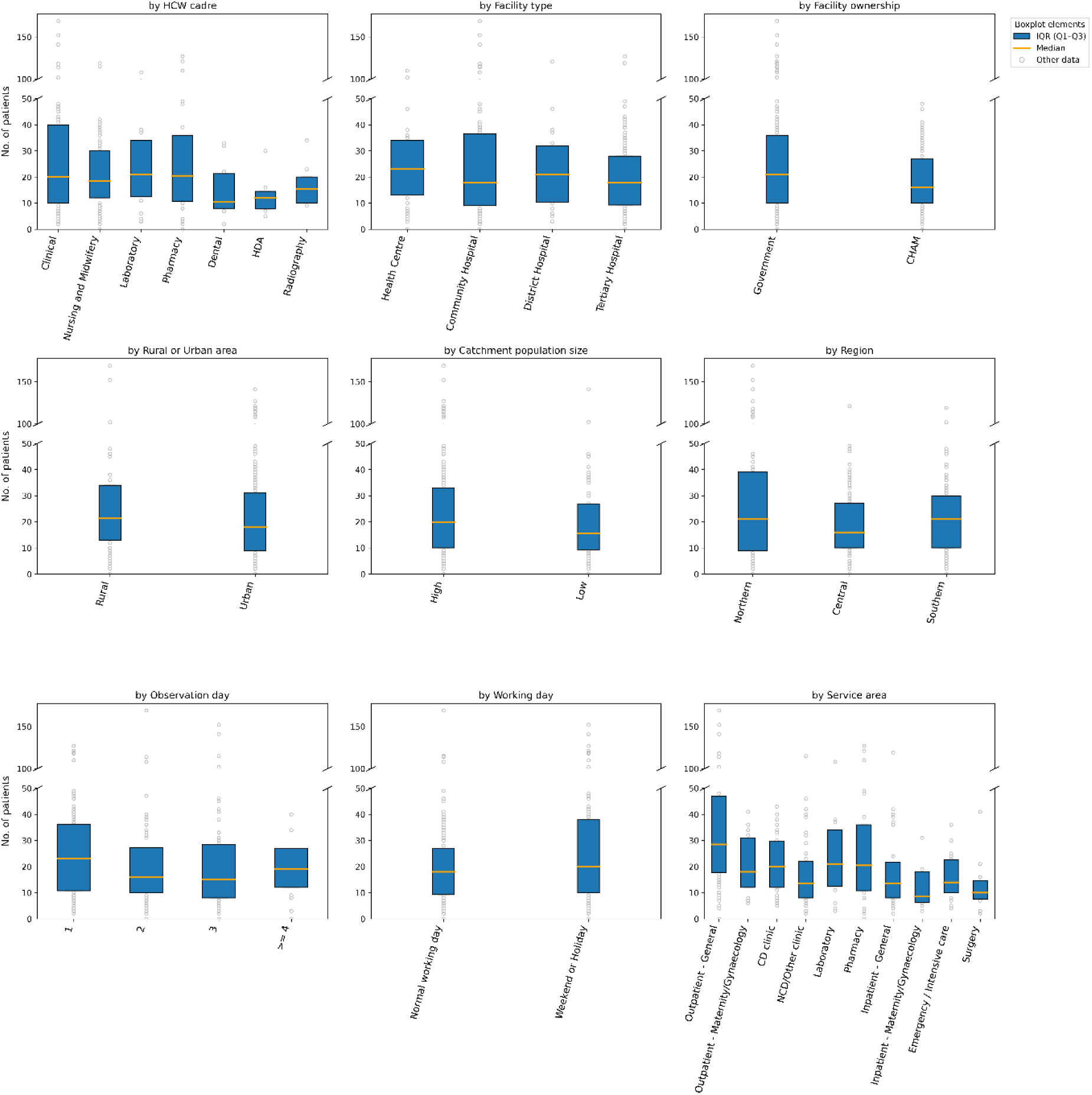
Daily patient loads: univariate analysis

#### A.2.4 Time per patient: univariate analysis

Figure A.4 shows the time that HCWs spent seeing a patient across different categories and characteristics. Due to the nature of services and patient conditions, the time per patient has substantial variations within each category as indicated by the IQRs and overall ranges. A surgery at a tertiary hospital could take more than 200 minutes of a clinical HCW time, whereas many activities such as sending the patient to do a malaria test and dispensing drugs could take less than 1 minute.

Significant differences in median values were also found between categories of most characteristics. Dental, HDA, and Radiography cadres had the highest time per patient with 8 to 11 minutes, followed by Nursing and Midwifery cadre with 5 minutes, Clinical and Laboratory cadres with 3 minutes, and Pharmacy cadre with 2 minutes. HCWs at Tertiary hospitals had spent 5 minutes per patient, higher than 3 minutes per patient at District hospitals, Community hospitals, and health centres. Government HCWs had spent 3 minutes per patient, lower than CHAM HCWs by 2 minutes. HCWs in Rural areas had spent 3 minutes per patient, lower than urban areas by 1 minute. HCWs in Northern region had spent 2 minutes per patient, lower than Central region by 2 minutes and Southern region by 3 minutes. Surgery clinics, Emergency care units, and Maternal/Gynaecological inpatient wards had the highest time per patient with 9 to 11 minutes, followed by CD clinics, Maternal/Gynaecological outpatient clinics, NCD/Other clinics, and General inpatient wards with 5 to 6 minutes, and Laboratories, General outpatient clinics, and Pharmacies with 2 to 3 minutes. Normal working days had a slightly higher time per patient than weekends or holidays by less than 1 minute.

**Figure A.4.**
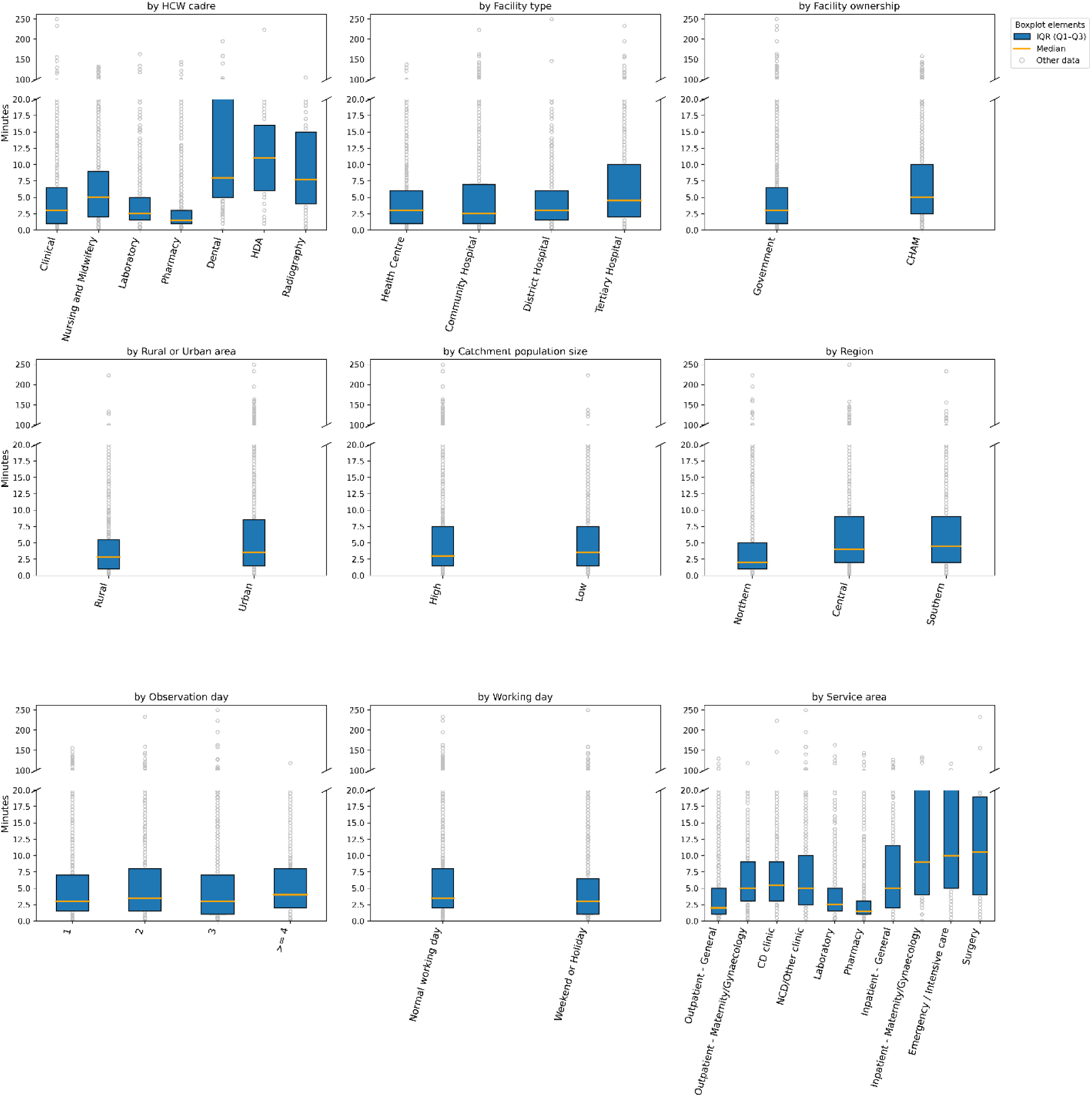
Time per patient: univariate analysis

#### A.2.5 Exemplar results on finer variable classification

Here we present the results for HCWs in Clinical and Nurse and Midwifery cadres and General and maternal/gynaecological outpatient clinics, with finer categorisations for HCWs and clinics. These groups are selected because they have more observations than others. The pairwise Mood’s Median tests results can be found in folder “Finer analysis - Pairwise Mood Median Test results” in the repository: 10.5281/zenodo.19852416.

For daily working time (Figure A.5) and daily patient loads (Figure A.7), we found no significant difference between three clinical sub-cadres and two nursing cadres. For daily working time allocation (Figure A.6), Medical Officer/Specialist had a significantly lower Unallocated time proportion than Clinical Officer/Technician and Medical Assistant by 12% to 14%. For time per patient (Figure A.8), Medical Officer/Specialist had a much higher value than Clinical Officer/Technician and Medical Assistant; with service area fixed, we further found that Clinical Officer/Technician had a slightly higher value than Medical Assistant in children outpatient clinics, and that Nurse and Midwifery Technician had higher values than Nurse Officer in antenatal and postnatal care clinics, whereas the latter had a higher value than the former in cervical cancer clinics. However, these differences should be carefully interpreted since the sample sizes for most cadre-location groups are very limited, as indicated in Figure A.7; in particular, there is only one day observation of one Medical Officer/Specialist in adult outpatient clinic.

**Figure A.5.**
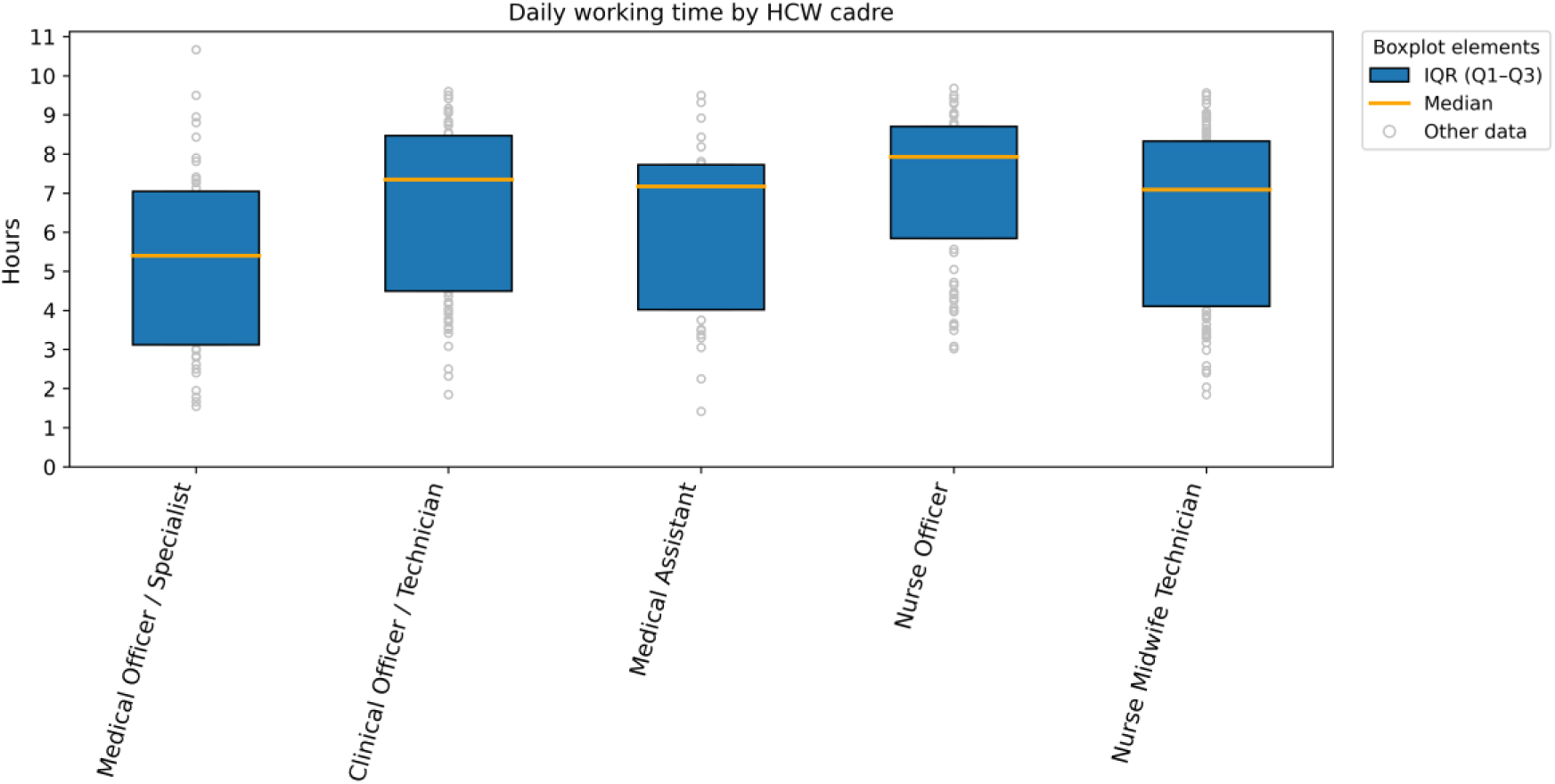
Daily working time: finer classifications for clinical and nursing staff

**Figure A.6.**
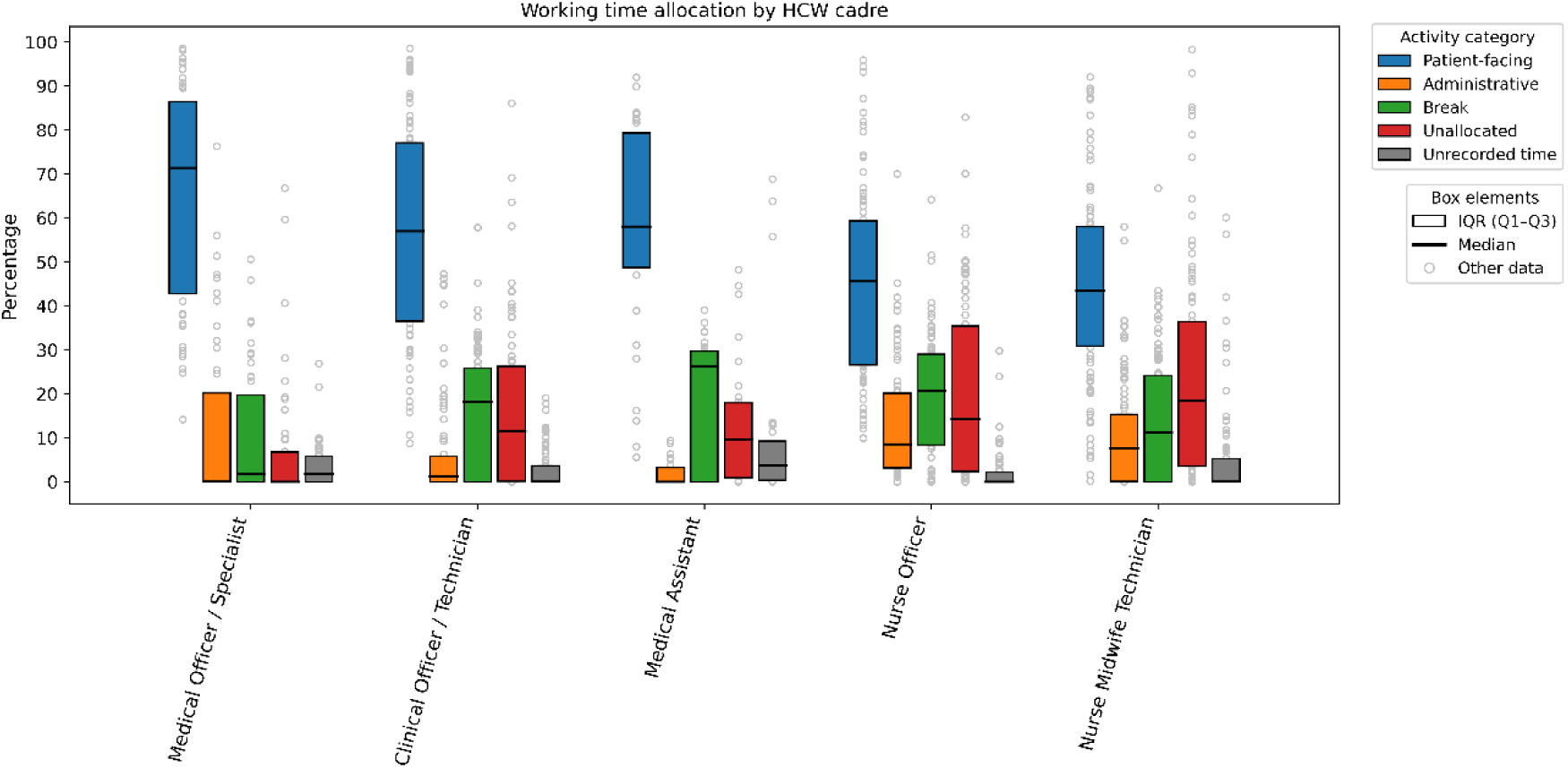
Daily working time allocation: finer classifications for clinical and nursing staff

**Figure A.7.**
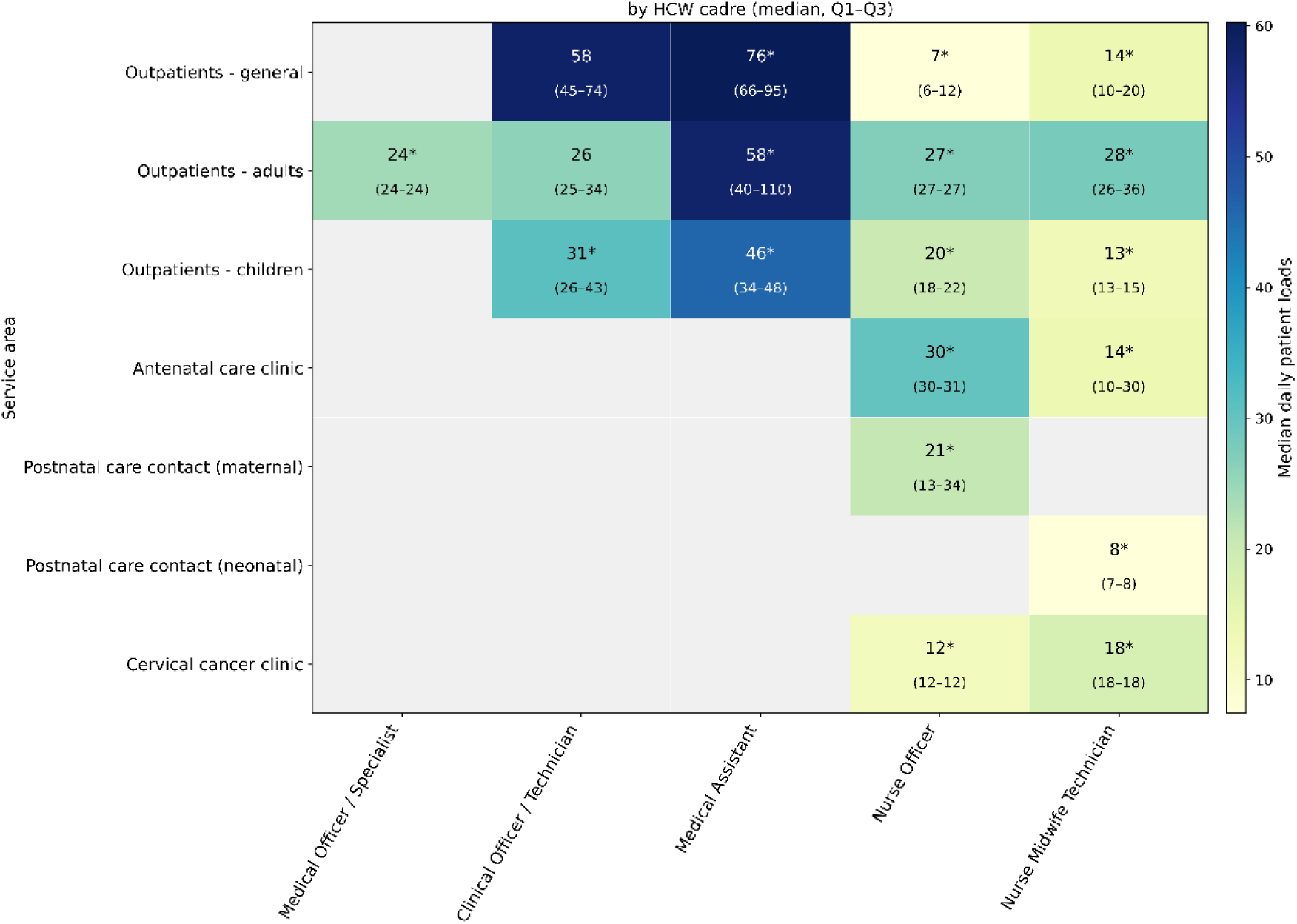
Daily patient loads: finer classifications for clinical and nursing staff and outpatient clinics

**Figure A.8.**
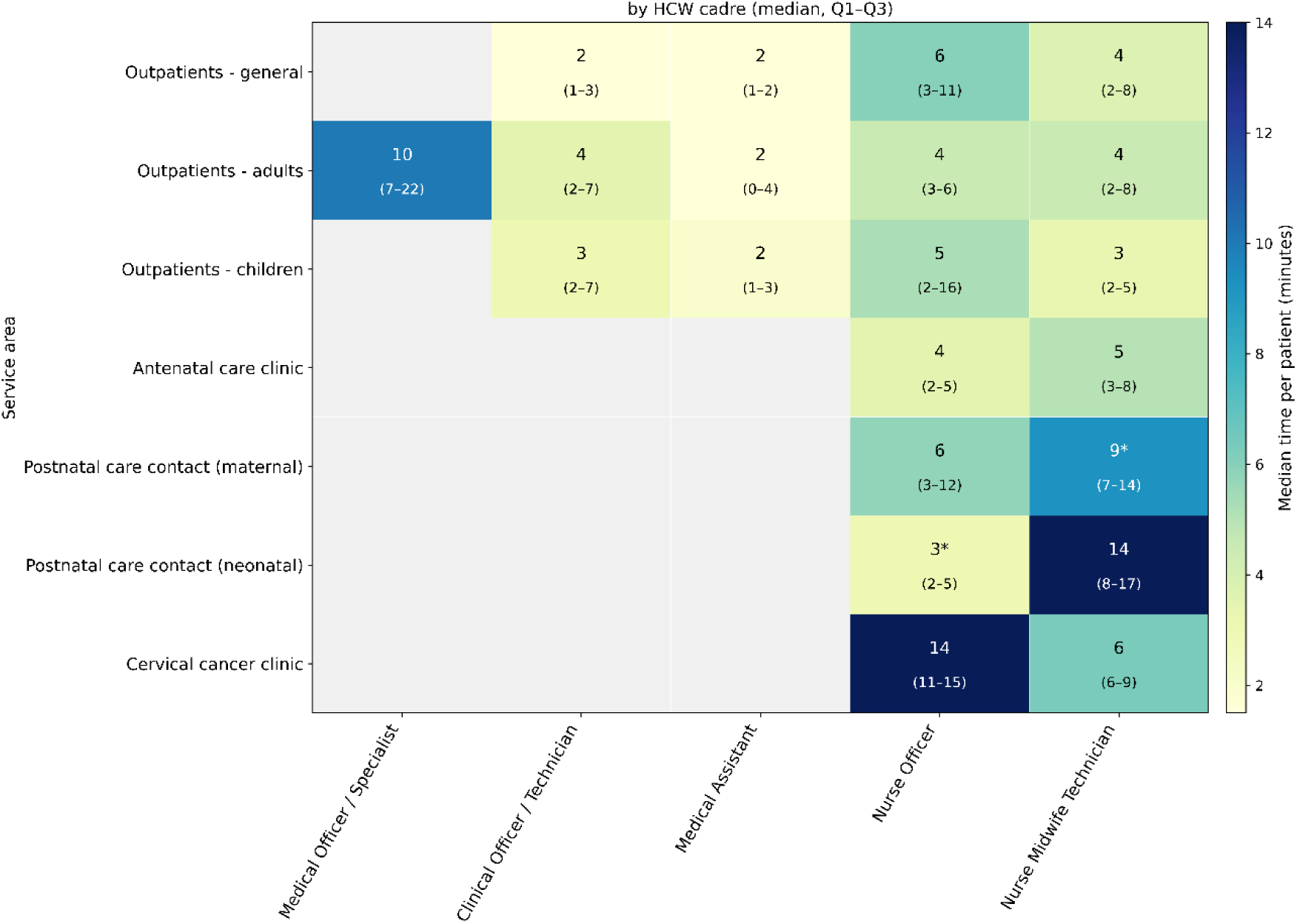
Time per patient: finer classifications for clinical and nursing staff and outpatient clinics

#### A.2.6 Temporal analysis on month

Below we present the HCW use patterns by month, noting that January and May had much fewer worker-day observations than other months (with 14 observations in January, 82 in February, 141 in March, 157 in April, 29 in May). As tested by pairwise Mood’s median tests, there is no significant difference across subgroups of months observed.

**Figure A.9.**
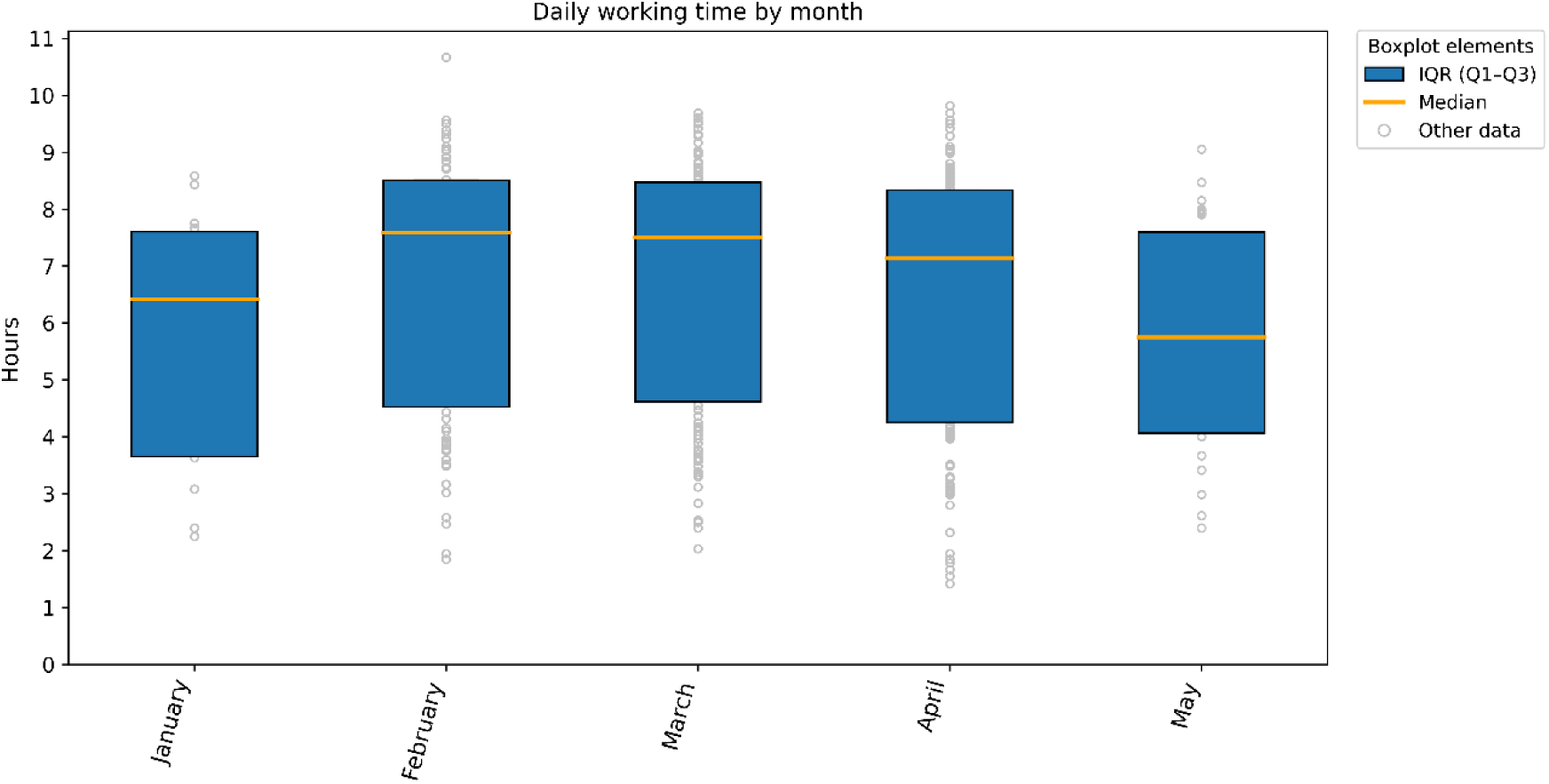
Daily working time by month

**Figure A.10.**
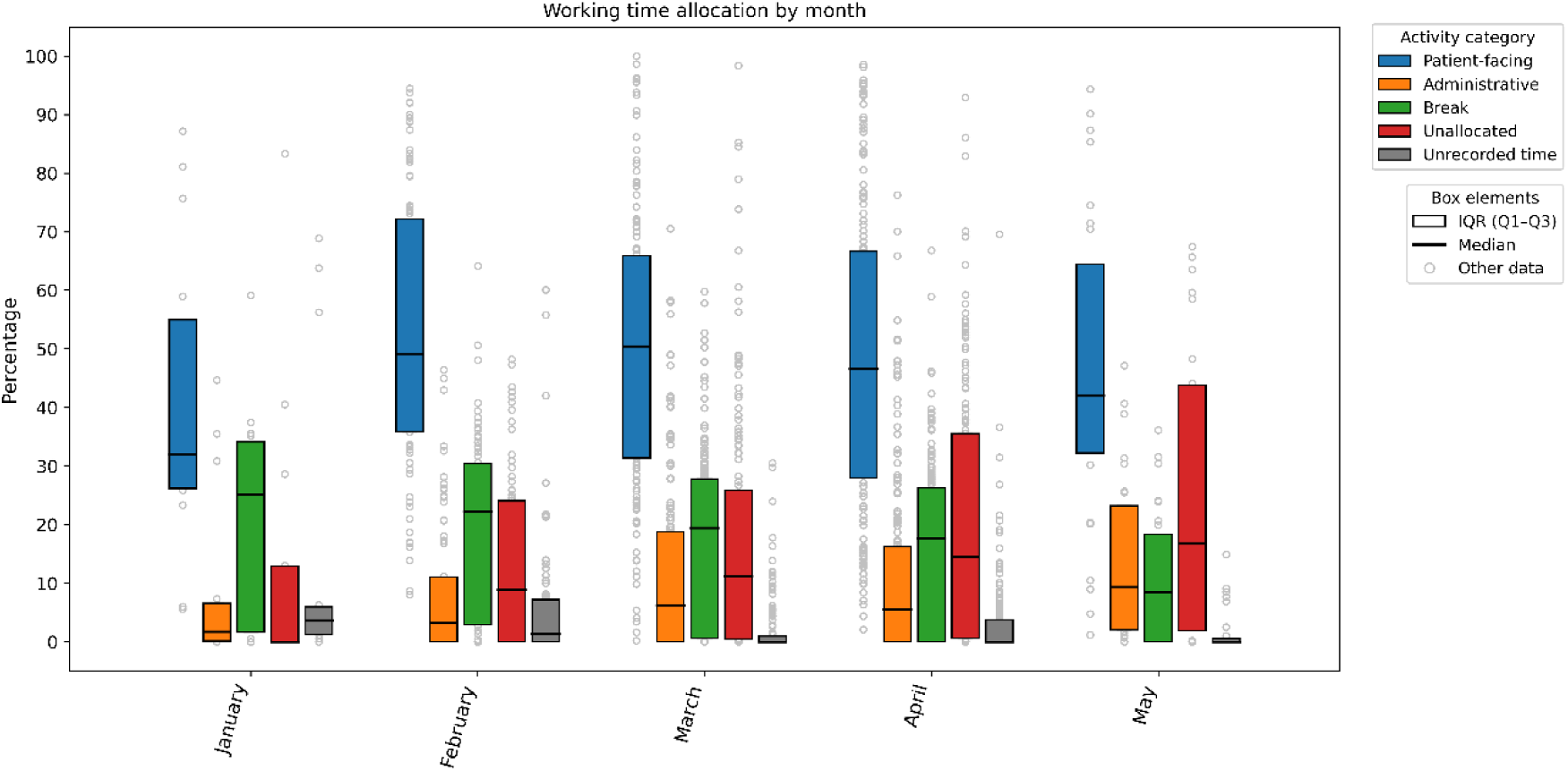
Daily working time allocation by month

**Figure A.11.**
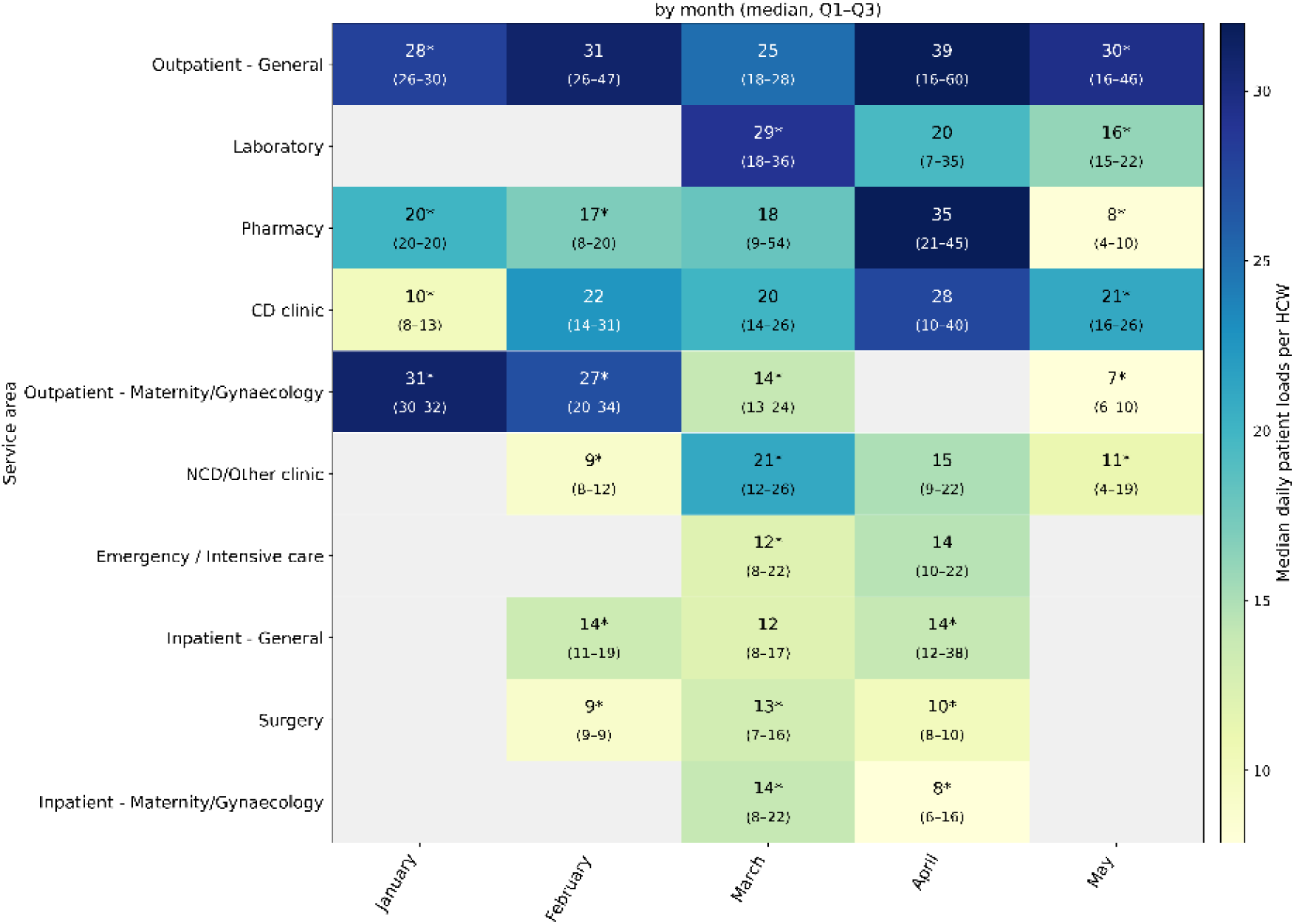
Daily patient loads by month

**Figure A.12.**
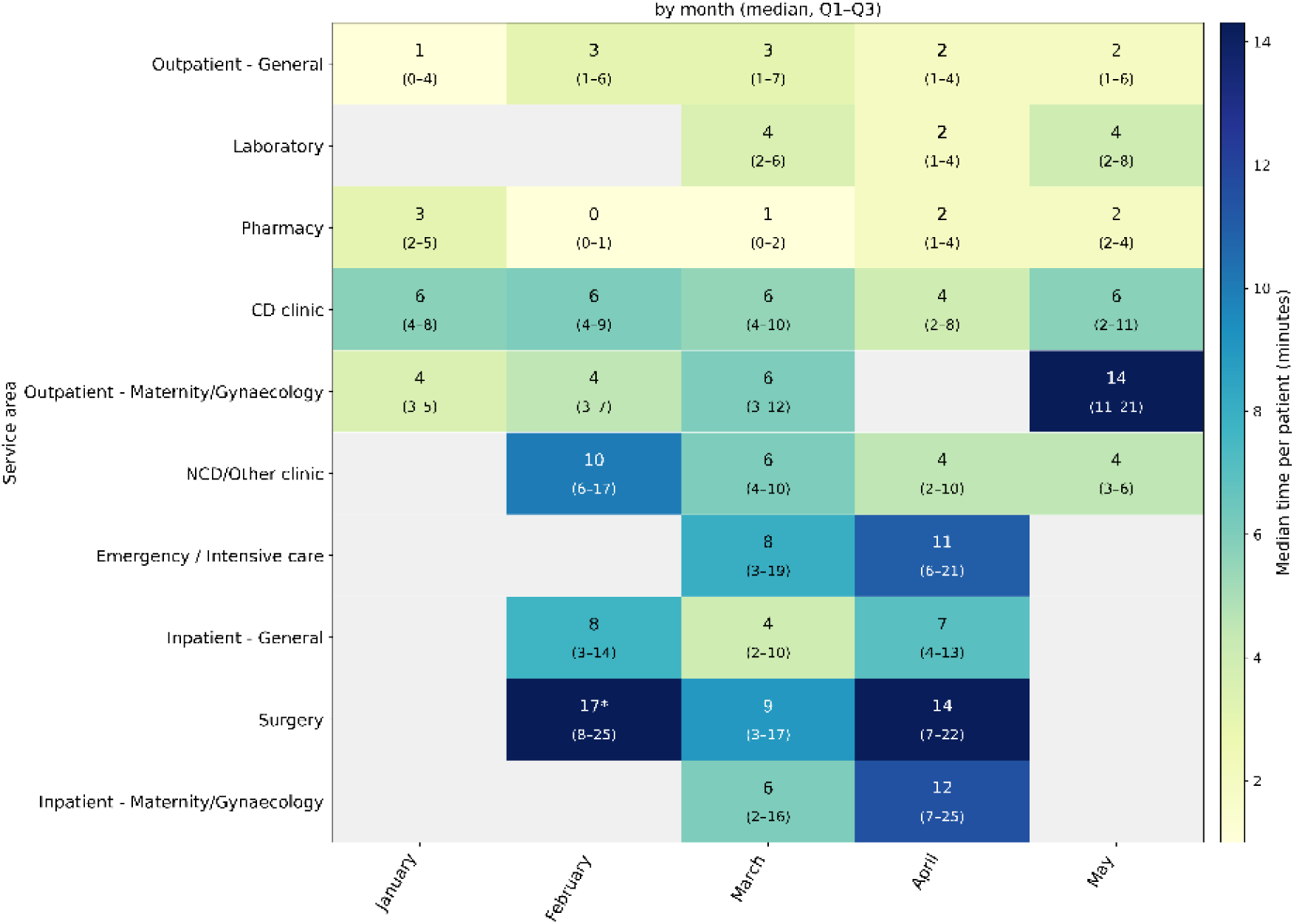
Time per patient by month

#### A.2.7 Overview results on mean and standard deviation

The HCWs worked a mean of 6.51 hours (SD 2.23) per day shift, among which Patient-facing time was 2.94 hours (SD 1.50; accounting for 48.86%, SD 24.49%, of the total working time), Administrative time was 0.83 hours (SD 1.13; 11.52%, SD 14.90%), Break time was 1.26 hours (SD 1.15; 16.98%, SD 14.75%), and Unallocated time was 1.22 hours (SD 1.41; 18.25%, SD 20.26%).

The mean number of patients seen per HCW per day was 29 (SD 28) across outpatient care areas, 17 (SD 18) across inpatient care areas, and 16 (SD 9) in emergency care, with the time per patient at 6.12 (SD 11.85) minutes, 12.68 (SD 19.98), and 15.46 (SD 16.97), respectively.

### A.3 Sensitivity analysis of data preprocessing

#### A.3.1 Daily working time allocation

The overall differences with the main analysis in daily working time allocation are that: a slightly, but not significantly lower Patient-facing time that was 2.62 hours (IQR 1.70-3.78), accounting for 43.52% (IQR 28.48%-61.67%) of the same total working time, and accordingly a minor extent of unrecorded time that was 0.23 hours (IQR: 0.07-0.47; 3.48%, IQR 1.23%-8%). In the main analysis, Patient-facing time was 2.82 hours (IQR 1.89-3.97), accounting for 47.52% (IQR 30.44%-66.87%) of total working time, whereas the unrecorded time gaps were negligible, with 0.00 hour (IQR 0.00-0.19; 0.00%, IQR 0.00%-3.79%).

Non-negligible unrecorded time proportions were found across characteristics (Figure A.13), with significant differences of median proportions between Clinical and Laboratory cadre, Health Centres and Hospitals, Rural and Urban areas, High and Low catchment population, and the Southern and Norther regions. These unrecorded time proportions correspond to slightly lower proportions of Patient-facing time per category of characteristics, which however show similar disparity patterns with the main analysis. See detailed results of pairwise Mood’s Median tests in folder “Sensitivity analysis - Pairwise Mood Median Test results” in the repository: 10.5281/zenodo.19852416.

**Figure A.13.**
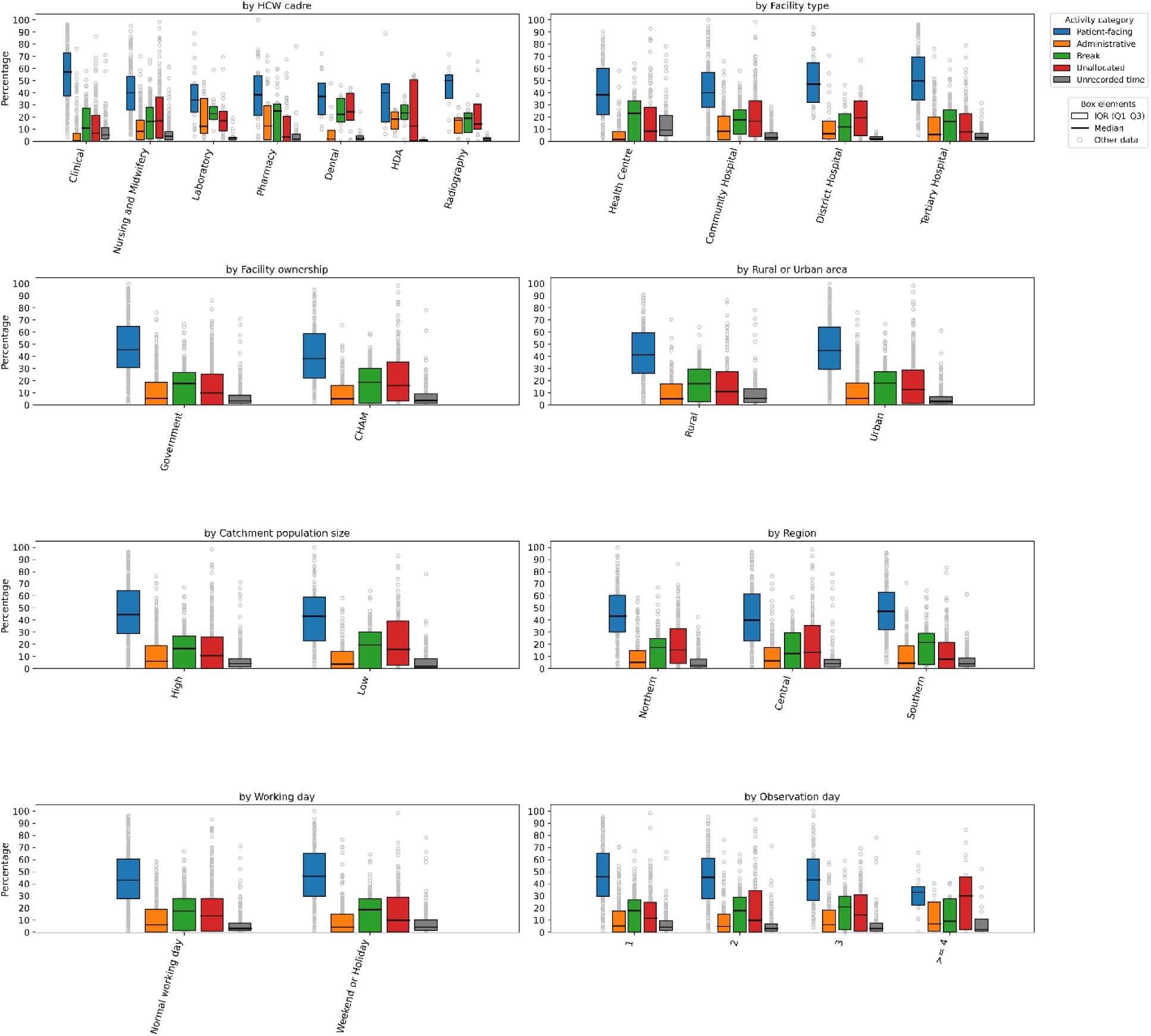
Daily working time allocation: Sensitivity analysis

**Figure A.14.**
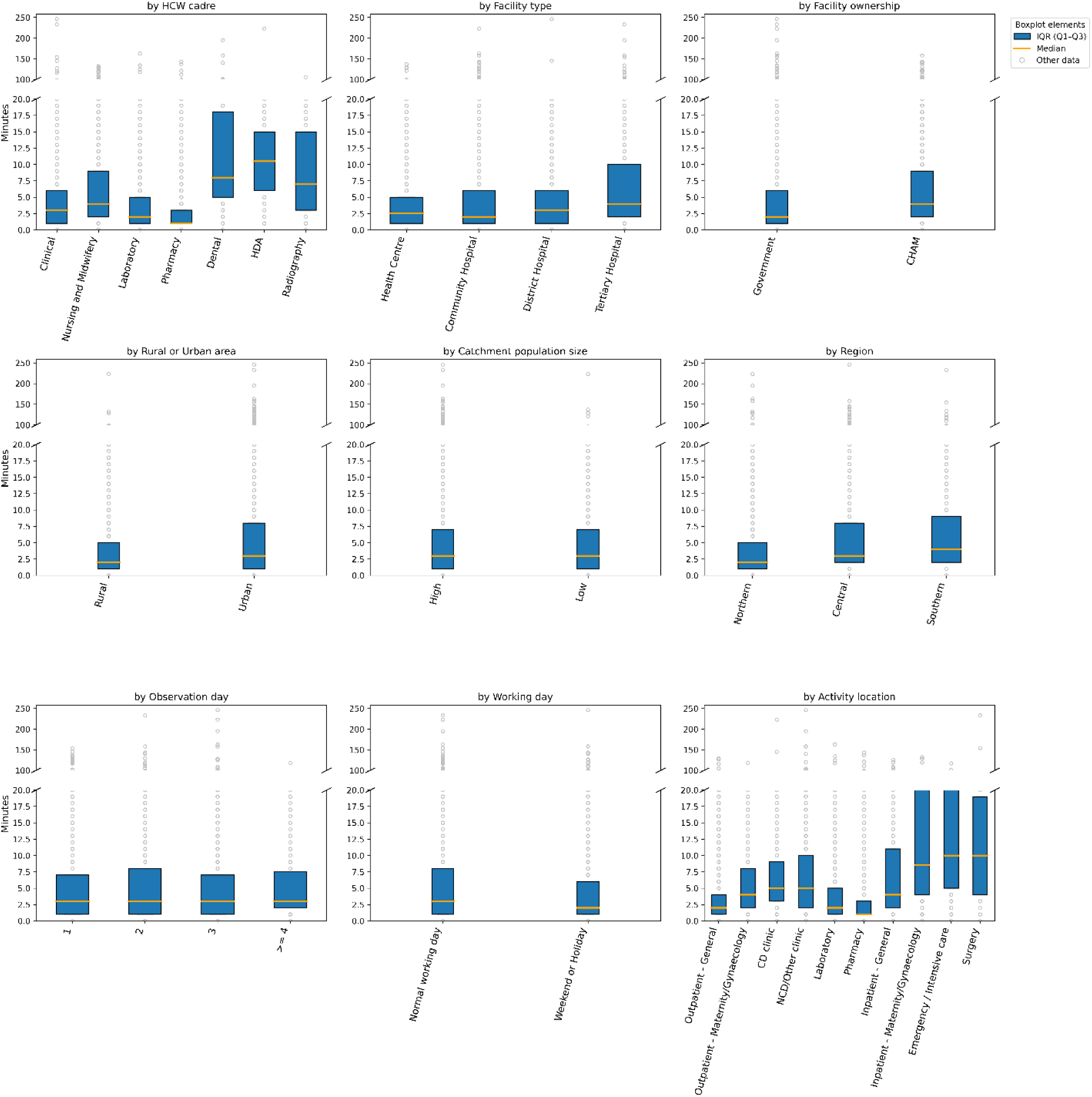
Time per patient: Sensitivity analysis

**Figure A.15.**
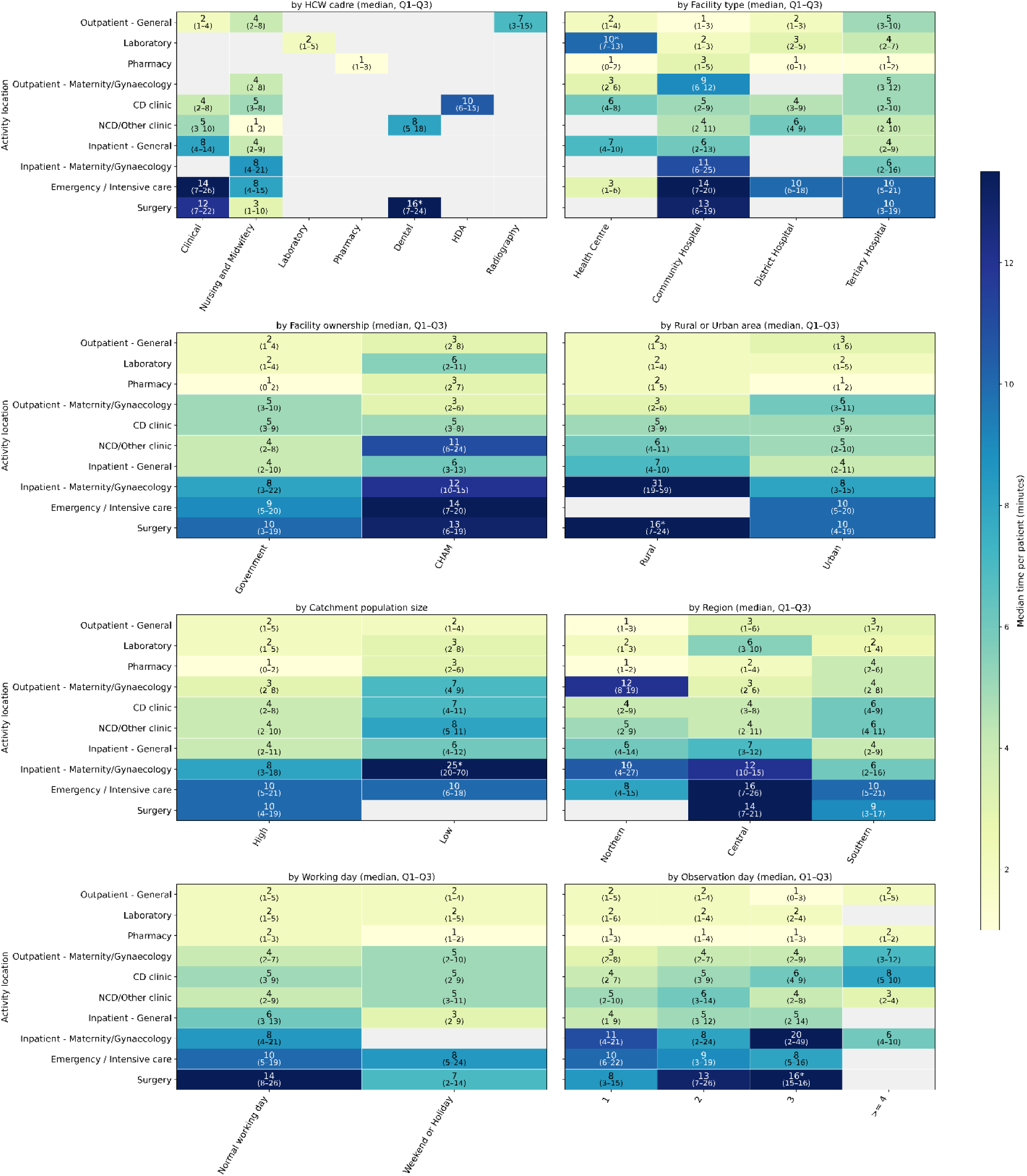
Time per patient per service area

#### A.3.2 Time per patient

The overall time per patient is consistent with the main analysis. Slight differences with the main analysis were found across categories of characteristics (Figure A.14): Clinical and Radiography cadres, both Government and CHAM facilities, Rural area, the Central region, Weekend/Holiday, General inpatient wards, and maternal/gynaecological outpatient clinics all saw 1-minute less per patient in median values, but with highly comparable IQRs. The disparities across categories of characteristics both with and without holding service area constant (Figures A.14 and A.15) are consistent with the main analysis (Figures A.4 and 4). See detailed results of pairwise Mood’s Median tests in folder “Sensitivity analysis - Pairwise Mood Median Test results” in the repository: 10.5281/zenodo.19852416.

## Reflexivity statement

The authors included 9 women and 12 men with professional experience in various areas of public health and health policy, spanning across multiple levels. This study has benefitted greatly from the deep expertise of the authors in Malawi and Sub-Saharan Africa, and their work across varied geographies within health system modelling and planning, public health research, and health policy making.

## Ethics statement

This research was approved by the College of Medicine Research and Ethics Committee (COMREC) in Malawi (Protocol Number: P.09/23-0297). All participants provided written informed consent prior to participation, and additional ethical safeguards were implemented to ensure confidentiality, voluntary participation, and minimization of risk.

## Acknowledgement

We greatly thank Dr Cosmas Kanyoma who has provided valuable advice on categorisation of HCW cadres, activities, and shift types with his expertise in this area and experience of working as a doctor in Malawi.

## Data Availability

The TMS data used for this study are publicly accessible in Thanzi La Mawa (TLM) datasets repository (https://github.com/HEPUMW/TLM-data-release/releases/tag/tlmdata_v1). In this data, we have removed or categorised variables that would pose risks to healthcare workers or patients being identifiable; those original variables cannot be shared due to confidentiality issues.

## Author contributions

BS conceived and designed the study, conducted data analysis, and drafted and revised the manuscript. WT and TC co-designed the study, supported with literature review, contributed to data interpretation, and revised the manuscript. PC and TC enabled access to data. BS, PC, JHC, WM, EM, PNM, VM, MS, DN, JM-B, TBH, WT, and TC supported with data curation. SB, TDM, SM (Mboma), SM (Mohan), MM, PNM, REM-W, AP, PR, MS, and TBH provided feedback and guidance on the manuscript. SM (Mboma), VM, DN, and JM-B provided expert input on health policy in Malawi and contributed to contextualisation of the study and data interpretation. The corresponding author (also the guarantor) BS accepts full responsibility for the work, had access to the data, and controlled the decision to publish. BS attests that all listed authors meet authorship criteria and that no others meeting the criteria have been omitted.

